# Wastewater-based Surveillance as a tool for monitoring and estimating COVID-19 incidence and trends: Insights from Germany, 2022-2024

**DOI:** 10.1101/2025.08.19.25333953

**Authors:** Susan Abunijela, Peter Pütz, Timo Greiner, Ann-Sophie Lehfeld, Alexander Schattschneider, Udo Buchholz, Jakob Schumacher

## Abstract

**Background:** Wastewater-based surveillance complements case-based surveillance systems by capturing pathogen signals shed in stool, enabling population-level monitoring independent of clinical testing. Its utility during the COVID-19 pandemic has been widely explored, but its responsiveness and interpretability relative to case-based systems remain insufficiently understood.

**Methods:** We analyzed data on COVID-19 or SARS-CoV-2 from July 2022 to December 2024 across 131 weeks, using wastewater surveillance and four case-based surveillance systems. These comprise syndromic surveillance systems at the population as well as the primary care level, and notification data, all aimed at monitoring COVID-19 incidence in Germany. We assessed agreement between wastewater viral load and disease incidence using visual inspection, cross-correlation analysis, and an estimated prevalence dynamic informed by a literature-based fecal shedding model. We derived retrospective translation factors and compared week-to-week trend directions between systems. Finally, we tested the predictive power of wastewater data using classification models to anticipate current week incidence trends.

**Results:** Wastewater SARS-CoV-2 viral load closely correlates with COVID-19 incidence trends from case-based systems, showing similar timing of peaks and troughs without notable time lags. Cross-correlation coefficients are highest with syndromic surveillance systems (up to 0.87) and lowest with notification data (0.43). Retrospective translation into incidence estimates works well on average, but week-to-week translation varies considerably. Wastewater-based models correctly predict the current week trend, as indicated by at least three of the four case-based systems, with about 68% probability.

**Conclusion:** Wastewater surveillance correlates well with COVID-19 incidence, but real-time translation to incidence lacks precision. Trend prediction for the current week may demonstrate improved accuracy and may be valuable when case reporting is limited or delayed.

## 1. Introduction

Disease surveillance systems, such as mandatory case-based reporting or syndromic surveillance, provide critical epidemiological data on disease incidence and severity. Wastewater-based surveillance (WBS) provides a complementary perspective that is independent of many factors that threaten consistency in case-based surveillance systems such as testing behavior [Wu et al., 2022]. By detecting and quantifying pathogen genome fragments excreted in stool and transported through the sewer system, wastewater data can monitor the infection burden at the population level, regardless of clinical testing or symptom status [Sosa-Hernández et al., 2022, Choi et al., 2020]. As a result, WBS has been adopted in many countries during the COVID-19 pandemic [Prado et al., 2021, Hart and Halden, 2020].

While the technical performance of WBS is well established, its integration into public health surveillance frameworks remains incomplete. WBS does not measure individual-level infections, and interpreting the quantitative signal from wastewater samples in terms of clinical incidence is challenging. Recent studies have attempted to bridge this gap using statistical and mechanistic models to link wastewater SARS-CoV-2 viral load with population-level incidence, prevalence, hospitalization rates, or the reproductive number [Mohring et al., 2024, McManus et al., 2023, Huisman et al., 2022, McMahan et al., 2021].

There is limited understanding of how quickly wastewater SARS-CoV-2 viral load reflects changes in disease dynamics compared to case-based surveillance systems, and whether it can reliably detect past and current trends in incidence. Although data from wastewater surveillance are sometimes interpreted qualitatively (e.g., “rising trend”), their quantitative translation into actionable indicators remains underdeveloped [McMahan et al., 2021].

This study addresses these gaps by evaluating the responsiveness, interpretability, and predictive potential of WBS during the COVID-19 pandemic in Germany. We compare SARS-CoV-2 WBS data with four established case-based surveillance systems that track COVID-19 incidence at the population level. Our goal is to assess how WBS performs in real-world settings and to determine its value as a complementary source of information.

We addressed the following questions:

1. How well does wastewater viral load correspond to disease incidence overall and particularly in terms of timeliness, increases, and decreases?
2. Can wastewater data be retrospectively translated into an estimate of disease incidence?
3. Do historical week-to-week trends from WBS agree with historical week-to-week trends in reported incidence?
4. Can wastewater data be used to predict disease incidence trends for the current week?

## 2. Data and Methods

### 2.1 Study period

The study period spanned from July 1, 2022, to December 31, 2024. During this period, data were collected for a total of 131 weeks from wastewater monitoring and four case-based surveillance systems that aimed to capture disease incidence. To smooth out noise in the measurement data, we applied generalized additive model for some analyses [Wood, 2017]. For all smoothed time series, we excluded the observations within the first and last five weeks of the study period, as smoothers tend to be less stable at the boundaries of time series due to the lack of neighboring data points to inform the smoothing process [Hastie and Tibshirani, 1987]. Therefore, analyses using smoothed time series include a total of 121 weeks.

### 2.2 Wastewater-Based Surveillance and Case-Based Surveillance Systems Utilized for COVID-19 Monitoring

WBS is a tool for detecting and monitoring pathogens circulating in wastewater [McMahan et al., 2021]. The German approach has been described in detail by Marquar et al. [2024] and Saravia et al. [2024]. In short, since July 2022, the German AMELAG project has monitored SARS-CoV-2 in wastewater, starting with 36 and increasing to 161 wastewater treatment plants (WWTPs) at the end of the study period. Wastewater samples at each site were usually collected twice per week at each WWTP using a 24-hour automatic sampler. Volume flow was used for normalization. At least two representative gene fragments (e.g. N1, N2, E, ORF, RdRp) were quantified by a network of 28 laboratories using digital PCR or quantitative real-time PCR. Geometric means of the measured gene fragments were aggregated weekly (Thursday to Wednesday) for each site, and then nationally across all WWTPs to obtain viral load values (gene copies/L). Weights were assigned based on population size in the catchment areas. An adjustment for mean differences in viral loads between different WWTPs and laboratories was applied. Further details are provided in Technical Guideline Part 4 for Wastewater Surveillance [AMELAG, 2025].

Wastewater data were compared with four case-based surveillance systems, three of which have been described previously [Loenenbach et al., 2024]. Brief descriptions of indicators derived from these systems are as follows:

1. German Notification System Incidence (GNS-I): This indicator is based on Germanýs mandatory notification system, which recorded PCR-confirmed SARS-CoV-2 infections reported by physicians, laboratories, and others to local public health authorities [Loenenbach et al., 2024, Dreesman and Benzler, 2005].
2. Participatory System Self-Reported Incidence (PS-SR-I): This indicator is based on the syndromic, participatory, population-based surveillance system *GrippeWeb*, in which participants voluntarily report weekly on the presence or absence of acute respiratory infections (ARI). Additionally, they can indicate whether their infection has been confirmed as SARS-CoV-2, for example, through a visit to a primary care provider or through self-testing at home [Loenenbach et al., 2024, Bayer et al., 2014].
3. Participatory System combined with Virological Positivity Rates Incidence (PS-VPR-I): This indicator combines ARI rates from *GrippeWeb* with SARS-CoV-2 positivity rates from the virological sentinel surveillance system, which collects respiratory samples from patients consulting primary care practices [Loenenbach et al., 2024, Buchholz et al., 2023].
4. Incidence of COVID-19 cases with medical consultation combined with the proportion of persons with ARI consulting a physician (Primary Care–COVID-ARI-I, PC-COVID-ARI-I): This indicator is derived from two data sources. First, the Sentinel for electronic recording of diagnostic codes for acute respiratory diseases (PC-ARI), which records ICD-coded COVID-19 diagnoses in primary care [Goerlitz et al., 2021]. Second, *GrippeWeb*, which provides the proportion of ill individuals with ARI who consult physicians. The population-level incidence rate (PC-COVID-ARI-I) is calculated by dividing the incidence of COVID-19 in primary care by the proportion of persons with ARI consulting a physician.

A more detailed description of these systems can be found in Table A1.

### 2.3 Question 1: How well does wastewater viral load correspond to disease incidence overall and particularly in terms of timeliness, increases, and decreases?

To address Question 1, we analyze the data using three approaches: visual comparison, cross-correlation analysis, and construction of a hypothetical prevalence curve using a weighted fecal shedding profile.

1. **Visual Inspection of Population Incidence and Wastewater Data**: We assess the resemblance between the course of wastewater viral load and disease incidence data by visually comparing smoothed wastewater viral load data with smoothed data from the four case-based surveillance systems described above. This comparison illustrates how closely wastewater data tracks population incidence.
2. **Cross-Correlation Analysis:** We quantify the degree of agreement and temporal relationship between wastewater viral load data and incidence data from each case-based surveillance systems by computing Pearson’s cross-correlation coefficients. This method measures the strength of the linear relationship between two time series and identifies the time lag that maximizes correlation. In other words, this analysis provides an overall measure of how closely wastewater data tracks disease incidence over time.
3. **Hypothetical Prevalence Curve Using a Weighted Fecal Shedding Curve:** We explore whether biological factors, such as fecal shedding dynamics, may influence the timing and shape of wastewater signals, particularly during increases and decreases in disease incidence. We construct a fecal shedding curve based on data reported in three published studies, each conducted at different stages of the COVID-19 pandemic and reflecting periods with different dominant SARS-CoV-2 variants (Table A2) [Wannigama et al., 2024, Arts et al., 2023, Wölfel et al., 2020]. Additional methodological details and results are provided in [Abunijela et al., 2025]. We apply exponential interpolation to convert measurements into daily values if data are only available on a coarser time scale. If data on viral shedding kinetics are available for different SARS-CoV-2 variants within a study, we calculate a median shedding pattern across variants, weighted by the number of cases reported for each variant and day. We standardize the data before aggregating them across the three studies. The aggregation is done by calculating the median viral shedding values for each day, weighted by the number of cases in each study. We then construct a hypothetical prevalence curve for each case-based surveillance system by combining incidence data with the derived fecal shedding pattern. Since the shedding pattern is estimated on a daily basis, we also convert weekly incidence data into daily values, for each incidence indicator, using exponential interpolation. Finally, we compare this hypothetical prevalence curve with wastewater viral load data.

### 2.4 Question 2: Can wastewater data be retrospectively translated into an estimate of disease incidence?

We calculate a weekly translation factor to convert smoothed wastewater viral load data into smoothed population incidence data, thereby enabling us to estimate the disease incidence based on wastewater data.

The translation factor for a time point *t* is generally calculated as:

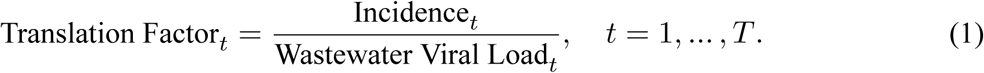

Thus, an average translation factor over time can be computed as:

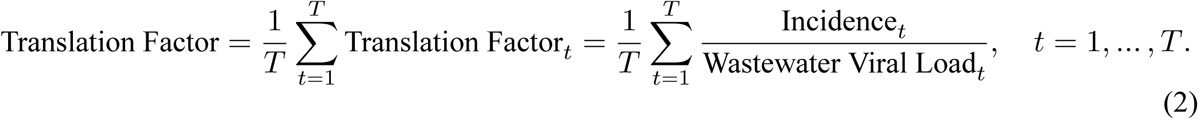

Non-contemporaneous versions of equations (1) and (2), i.e. considering lags between wastewater viral load and incidence, could be calculated accordingly. In this study, we solely consider contemporaneous translation factors.

This translation is considered retrospective because the smoothing procedure relies on “future data” (i.e. data that become available after each time point considered) which is, in practice, not available in real-time [Hastie and Tibshirani, 1987]. Therefore, we obtain a retrospective estimate of how wastewater viral load might correspond to disease incidence at a given point in time. We plot the translation factor over time.

To estimate an average retrospective translation factor, we plot one-to-one comparisons between wastewater viral load and each of the four incidence indicators in a scatter plot so that one week reflects one point in the plot. By applying regression through the origin (anchoring the line at 0/0), we derive the slope, which allows us to translate an increase in viral load into an estimate of an increase in population incidence.

### 2.5 Question 3: Do historical week-to-week trends in wastewater agree with historical week-to-week trends in reported incidence?

To assess if trends in wastewater data agree with trends in the other four surveillance systems, percentage changes in smoothed weekly data are calculated. This approach allows us to evaluate temporal coherence between systems beyond absolute levels.

We compute Pearson’s cross-correlations between week-to-week percentage changes, i.e. changes from one week to the previous one, in wastewater viral load and each incidence system to estimate the strength of linear associations over time.

To further interpret these changes as trends, we grouped them into three categories for each system:

- Increase (change compared to previous week > 5%)
- Decrease (change compared to previous week < -5%)
- Stable (change compared to previous week between -5% and 5%)

For the four case-based surveillance systems, we summarized these trend categories by using an indicator that marks whether at least three out of four systems fall into the same category. To this end, the case-based surveillance systems together can indicate “Increase”, “Decrease” or “Stable” if at least three out of the four systems have the same respective trend or “Unclear” if less than three systems show the same trend. This joint indicator derived from the case-based surveillance systems can be considered as a benchmark for the unobserved true trend in disease incidence. Agreement of this indicator with wastewater viral load is then given if both result in the same week-to-week trend category.

As described above, the smoothing method applied relies for each week on data from future weeks. Thus, the degree of agreement measured here reflects a retrospective assessment.

### 2.6 Question 4: Can wastewater data be used to predict disease incidence trends for the current week?

In real-life, the incidence systems may experience delays, data gaps, or reduced reliability. Assuming that only wastewater monitoring data is available, we try to predict the incidence trends for the current week using wastewater data up to the current week. To assess the predictive power, the target variable and benchmark in this analysis is a joint disease trend indicator derived from the four case-based surveillance systems, as described in the methods section for Question 3. We grouped this variable into three categories:

- Increase
- Decrease
- Other (a combination of “Stable” and “Unclear” trends to reduce class imbalance and ensure sufficient sample sizes per class).

We train three machine learning models aimed at classifying this target variable, each of the three models utilizes the following set of features (input variables):

- Trends derived from a 2-week moving average of viral loads in wastewater data, calculated by averaging the viral loads from the current and preceding week. For example, the trend for week 6 is computed by averaging the viral loads from weeks 5 and 6 and comparing this to the average from weeks 4 and 5 as described above in Question 3.
- Their first and second lags. In the example above, the first lag is computed by the trend comparing the average viral loads from weeks 4 and 5 to those from weeks 3 and 4. The second lag is computed analogously.
- Raw weekly viral loads in wastewater.
- Their first and second lags. The first lag corresponds to the raw viral load from the previous week (e.g., week 5 when predicting week 6). The second lag corresponds to the viral load from two weeks earlier (e.g., week 4 when predicting week 6).

We apply a multinomial regression with L1-Penalty (LASSO = least absolute shrinkage and selection operator), a support vector machine, and a random forest. All models are trained and evaluated using a nested blocked cross-validation procedure [Varma and Simon, 2006]. For the inner loop, we apply a 10-fold blocked cross-validation to tune hyperparameters and select the best-performing model based on overall classification accuracy. For the outer loop, we apply another 10-fold blocked cross-validation to estimate the model’s generalization error. Nested blocked cross-validation allows us to use the entire dataset for both training and testing, which is important given our small sample size, and is appropriate for time series [Bergmeir et al., 2018, 2014, Bergmeir and Benítez, 2012]. Non-dependent cross-validation is used in both loops to avoid overlapping in time between training and test data sets, which is essential for avoiding information leakage in time-dependent data [Bergmeir et al., 2018]. Finally, we select the model with the highest mean overall accuracy in the inner loop. Its performance is then evaluated in the outer loop in terms of the overall accuracy as well as on the positive predictive value and the sensitivity for the trend categories “Increase” and “Decrease”.

## 3. Results

### 3.1 Question 1: How well does wastewater viral load correspond to disease incidence overall and particularly in terms of timeliness, increases, and decreases?

#### 3.1.1 Visual inspection of population incidence and wastewater data

Visual inspection of the raw and smoothed data indicates that wastewater viral load mirrors the timing of rises and falls in COVID-19 incidence closely (Figure 1).While there are differences in the height of peaks across surveillance systems, peaks and troughs in wastewater data broadly coincide with those observed in the participatory syndromic systems (PS-SR-I, PS-VPR-I) and the sentinel system (PC-COVID-ARI-I). An exception is the GNS-I, which shows a consistently diverging trajectory with lower incidence values starting in late 2022.

**Figure 1:**
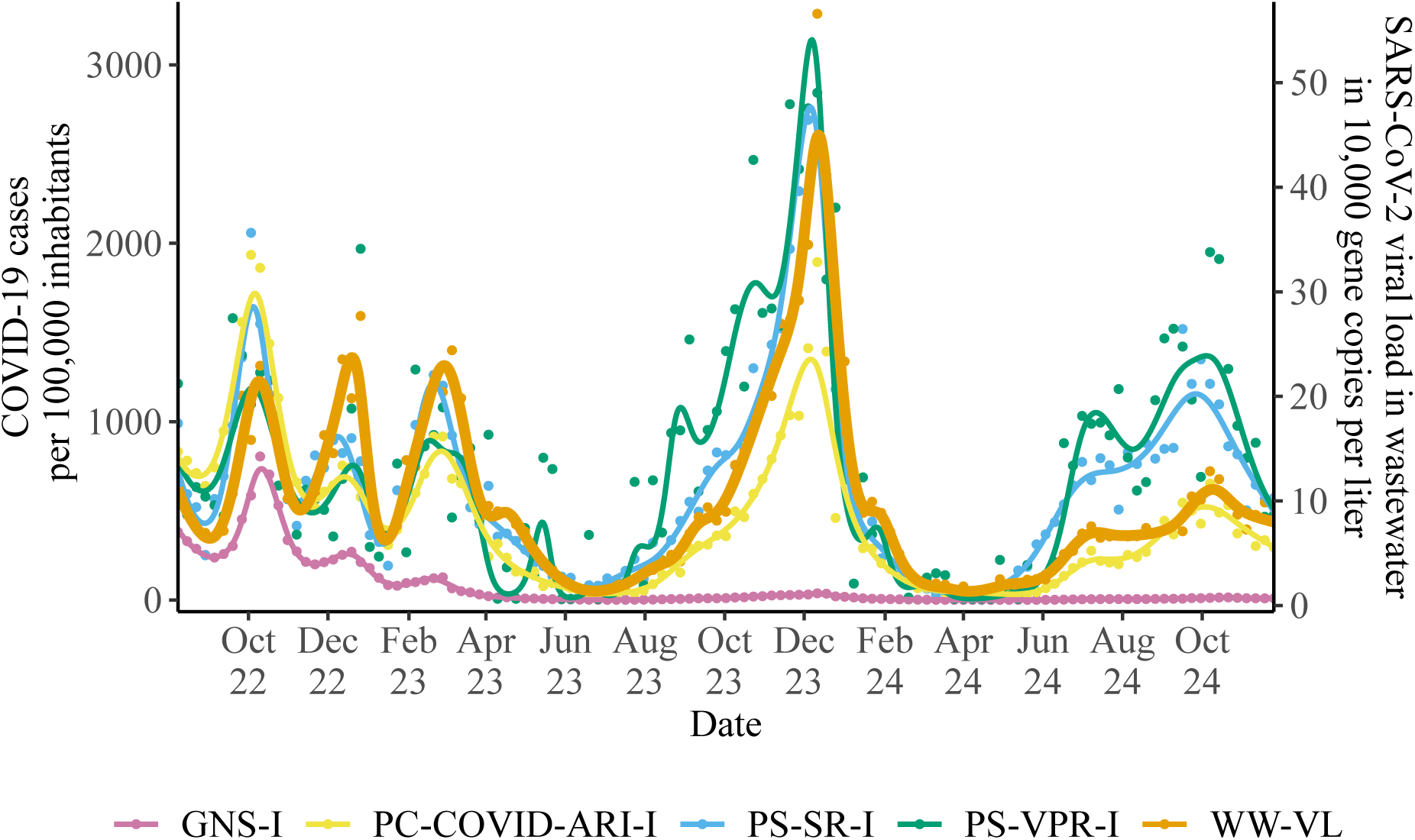
Comparison of wastewater SARS-CoV-2 viral load with COVID-19 incidence trends from cased-based surveillance systems, highlighting temporal agreement and differences in peak magnitudes. GNS-I = German Notification System of COVID-19, PC-COVID-ARI-I = Sentinel for Electronic Recording of Diagnoses of Acute Respiratory Infections, combined with Participatory System, PS-SR-I = Participatory System Self-Reported, PS-VPR-I = Participatory System Virological Positivity Rate, WW-VL = Wastewater Viral Load.

The variability of data around the smoothed curves differed among systems, with PS-VPR-I exhibiting the largest fluctuations. Even GNS-I also agrees with the peaks of the first three COVID-19 waves depicted before values become very low. Thus, the overall patterns of wastewater data and the other systems are quite consistent. In particular, during periods of rising incidence, wastewater viral load and case-based curves show largely synchronous behavior, with no consistent advantage in timeliness for any system. At times, the slope of the rise in the wastewater viral load is somewhat steeper, such as during the second (Dec 2023) and third (Feb 2023) waves, but not during the fourth (Oct–Dec 2023) and fifth (Jun–Aug 2024) waves (Figure 1).

#### 3.1.2 Cross-correlation analysis of incidence and wastewater data

The strongest cross-correlation of wastewater viral load is observed with PS-SR-I (correlation coefficient: 0.87), followed by PC-COVID-ARI-I (0.83) and PS-VPR-I (0.73) (Figure 2). The weakest cross-correlation was found with notification data (GNS-I, 0.43). The peak correlation for PS-SR-I and PS-VPR-I is observed when wastewater data are shifted back by one week, respectively, suggesting that wastewater data lag slightly behind incidence trends. However, the contemporaneous correlation, i.e. when no time shift was applied, is almost identical. For GNS-I and PC-COVID-ARI-I, the peak correlation is captured when no time shift is applied. These findings suggest that trajectories in wastewater data and incidence of COVID-19 exhibit no relevant time shift, supporting the visual impressions from Figure 1.

**Figure 2:**
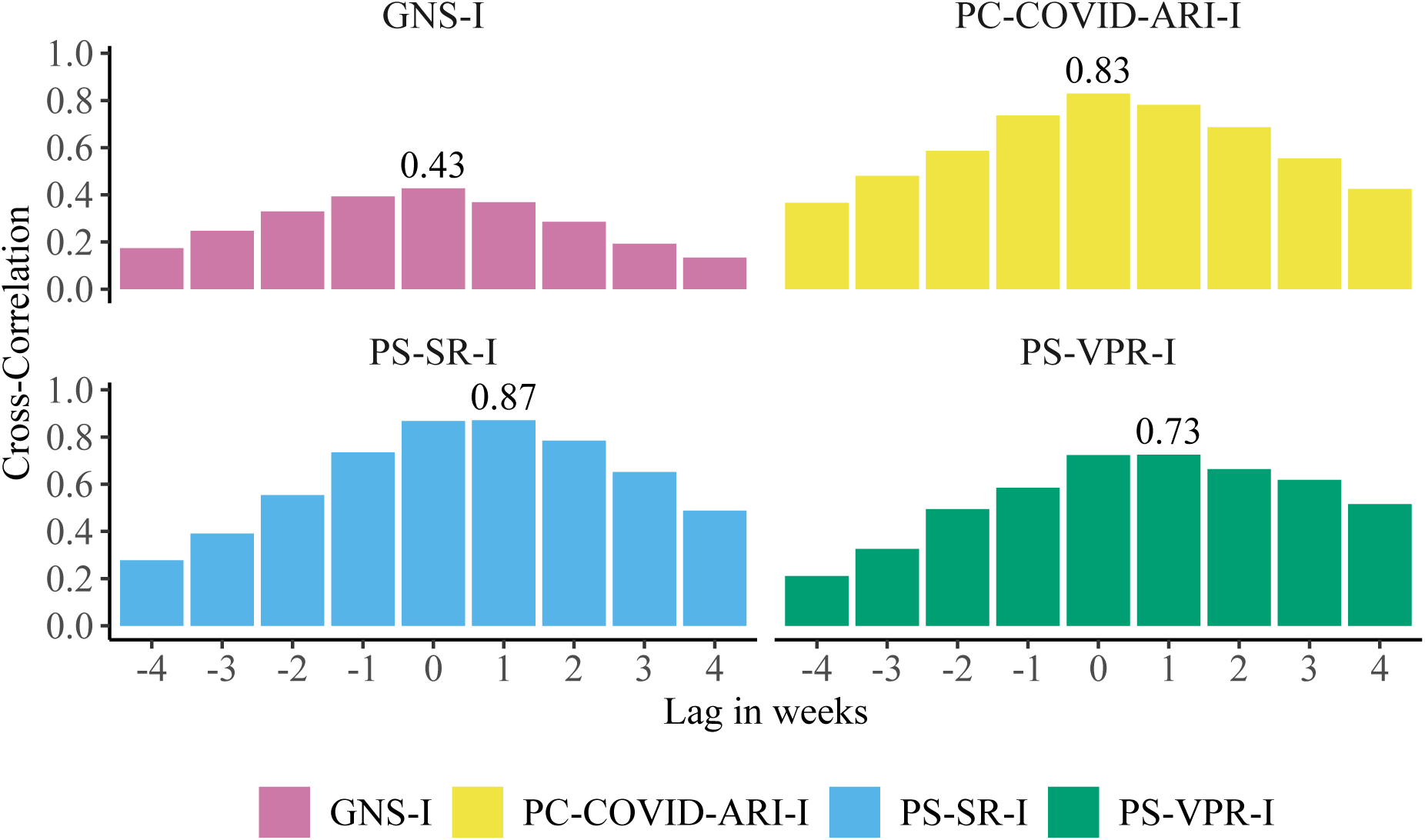
Cross-correlation between wastewater SARS-CoV-2 viral load and four COVID-19 incidence indicators at varying lags in weeks. A correlation for a lag of +2 weeks, e.g., indicates the correlation between current incidence and wastewater viral load measured two weeks back in time. GNS-I = German Notification System, PC-COVID-ARI-I = Sentinel for Electronic Recording of Diagnoses of Acute Respiratory Infections, combined with Participatory System, PS-SR-I = Participatory System Self-Reported, PS-VPR-I = Participatory System Virological Positivity Rate.

#### 3.1.3 Hypothetical prevalence curve using a fecal shedding curve

Figure 3 shows the normalized fecal shedding curve of SARS-CoV-2, derived from data reported in three published studies [Wannigama et al., 2024, Arts et al., 2023, Wölfel et al., 2020]. The aggregated curve displays a clear rise in viral shedding beginning around 5 days after symptom onset, followed by a gradual decline starting at around day 10.

**Figure 3:**
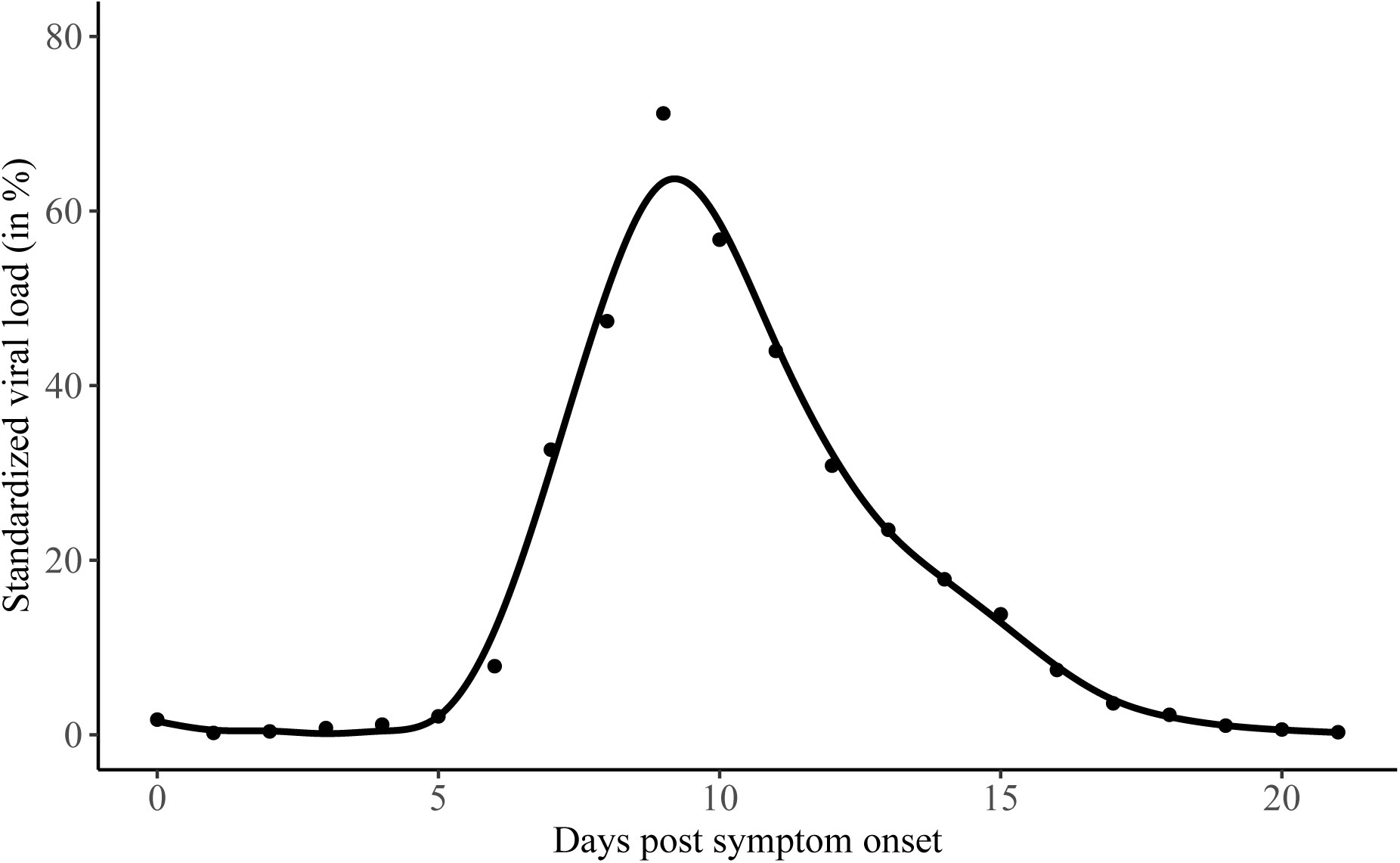
Fecal shedding distribution of SARS-CoV-2 after symptom onset. Measurements from three studies [Wannigama et al., 2024, Arts et al., 2023, Wölfel et al., 2020] are exponentially interpolated to obtain daily data and then standardized. Depicted are the median daily standardized values of the three studies, weighted by the number of cases per study, and smoothed values obtained by using a generalized additive model.

This pattern indicates that fecal shedding at peak magnitude is not sustained over several weeks. Instead, the viral load in stool peaks within a short time frame after symptom onset and declines relatively quickly. Figure 4 depicts this relationship more clearly: When deriving a hypothetical prevalence curve from PS-SR incidences and our fecal shedding curve, both, the resulting curve and the wastewater viral load coincide narrowly with the incidence curve. In particular, the steep decline of the incidence in January 2024 is accompanied by a decline just as steep of both the hypothetical prevalence and the wastewater curve. Corresponding figures for the three other case-based surveillance systems exhibit similar patterns (Figures A1, A2, A3). Moreover, cross-correlations between the derived hypothetical prevalence curves and wastewater viral load are very similar to the cross-correlations between the original incidence curves and wastewater viral load from Figure 2, albeit a bit shifted due to the lag introduced by the duration of the fecal shedding process (Figure A4).

**Figure 4:**
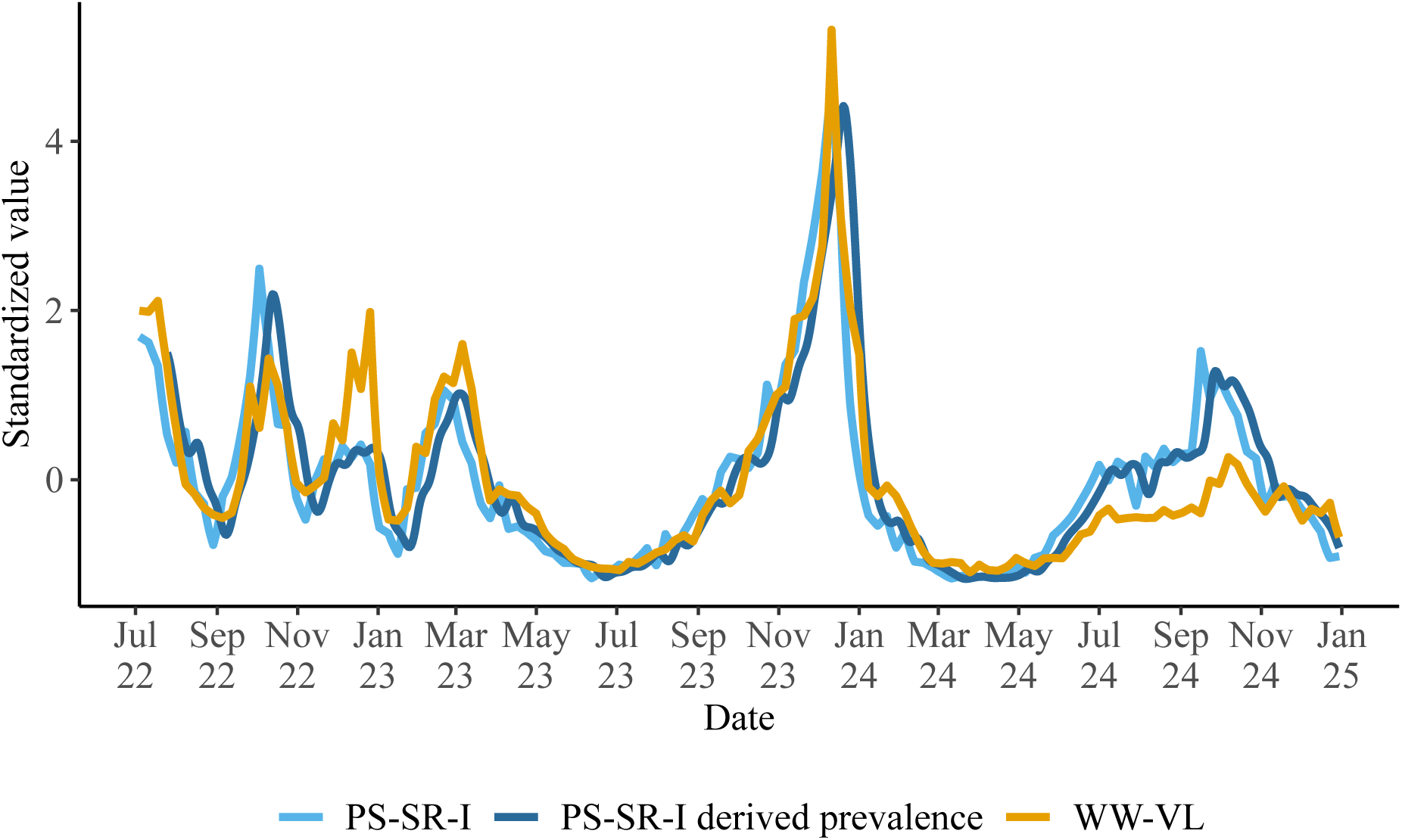
Hypothetical prevalence for PS-SR-I. PS-SR-I incidences were multiplied with the fecal shedding distribution shown in Figure 3 to derive a hypothetical prevalence curve for PS-SR-I. Shown are standardized values for the resulting hypothetical prevalence, the original incidence for PS-SR-I and the viral load in wastewater. PS-SR-I = Participatory System Self-Reported, WW-VL = Wastewater Viral Load.

### 3.2 Question 2: Can wastewater data be retrospectively translated into an estimate of disease incidence?

The left panel of Figure 5 shows the weekly (contemporaneous) translation factors for the case-based surveillance systems over time. The unit of the translation factor is the incidence of COVID-19 cases per 100,000 inhabitants divided by 10,000 RNA copies of SARS-CoV-2 per liter of wastewater. Thus, a translation factor of 100, for instance, means that 10,000 RNA copies/L wastewater correspond to 100 cases per 100,000 inhabitants per week.

**Figure 5:**
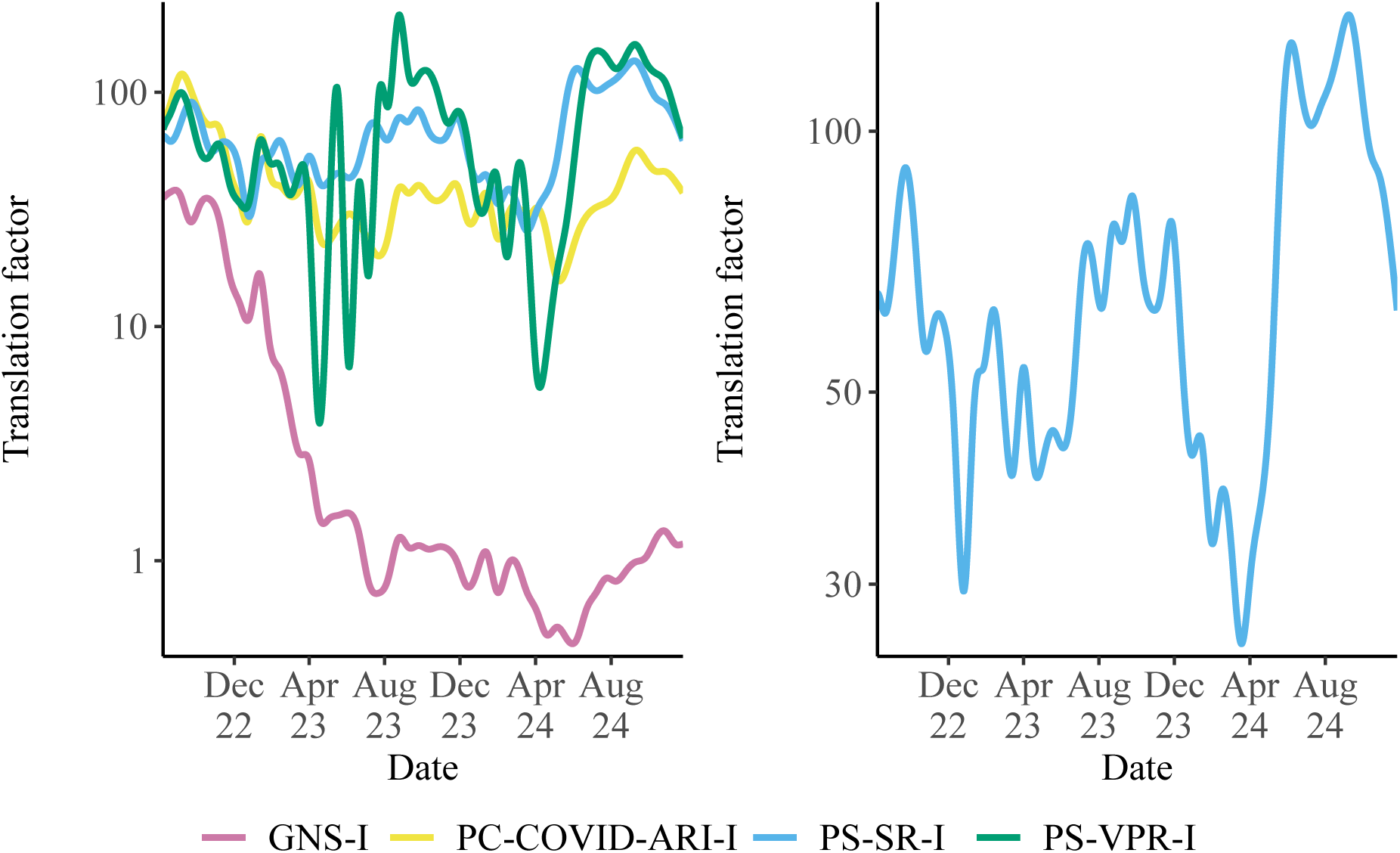
Translation factors (incidence per 100,000 inhabitants per 10,000 RNA copies SARS-CoV-2 per L wastewater) over time, plotted on a log10 scale. *Left panel*: Weekly translation factors for GNS-I, PC-COVID-ARI-I, PS-SR-I, and PS-VPR-I. *Right panel*: Zoomed-in view of weekly translation factors for PS-SR-I. GNS-I = German Notification System of COVID-19, PC-COVID-ARI-I = Sentinel for Electronic Recording of Diagnoses of Acute Respiratory Infections, combined with Participatory System, PS-SR-I = Participatory System Self-Reported, PS-VPR-I = Participatory System Virological Positivity Rate.

Among the systems, the translation factor for PS-SR-I exhibits the most stable values, generally ranging between 25 and 135 (zoomed in on the right panel of Figure 5). While relatively consistent during some periods, such as between mid-2023 and end-2023, the translation factor fluctuates increasingly in other periods, especially from 2024 onward. A comparison with Figure 1 shows that exceptionally high or low translation factors commonly - but not exclusively - occur in periods of low wastewater viral load values. Variations in the translation factor over time are even more evident for the other surveillance systems, with the notification system (GNS-I) generally showing a downward trend. Overall, the panels indicate that the translation factors remain relatively stable only over short time intervals and vary considerably across systems and time periods.

When omitting the time dimension, by fitting a linear regression through the origin, one can see that the regression slopes range from 9.04 COVID-19 cases per 100,000 inhabitants per 10,000 RNA copies SARS-CoV-2 (GNS-I) to 64.37 (PS-VPR-I) (Figure 6). The two mainly participatory system based indicators (PS-SR-I and PS-VPR-I) show the highest and very similar slopes (60.47 and 64.37, respectively). This indicates that an increase of the SARS-CoV-2 RNA concentration in wastewater corresponds to a larger increase in reported incidence in these two indicators compared to the other two incidence indicators. The notification-based system (GNS-I) is associated with the lowest slope (9.04). The slope for PC-COVID-ARI-I, the primary care-based indicator, lies in the mid-range at 42.52, but is closer to the PS-based indicators. The coefficients of determination (R²), that indicate the goodness-of-fit of the regression curves, also show notable differences, ranging from 0.36 for the notification data (GNS-I) to 0.89 for PS-SR-I.

**Figure 6:**
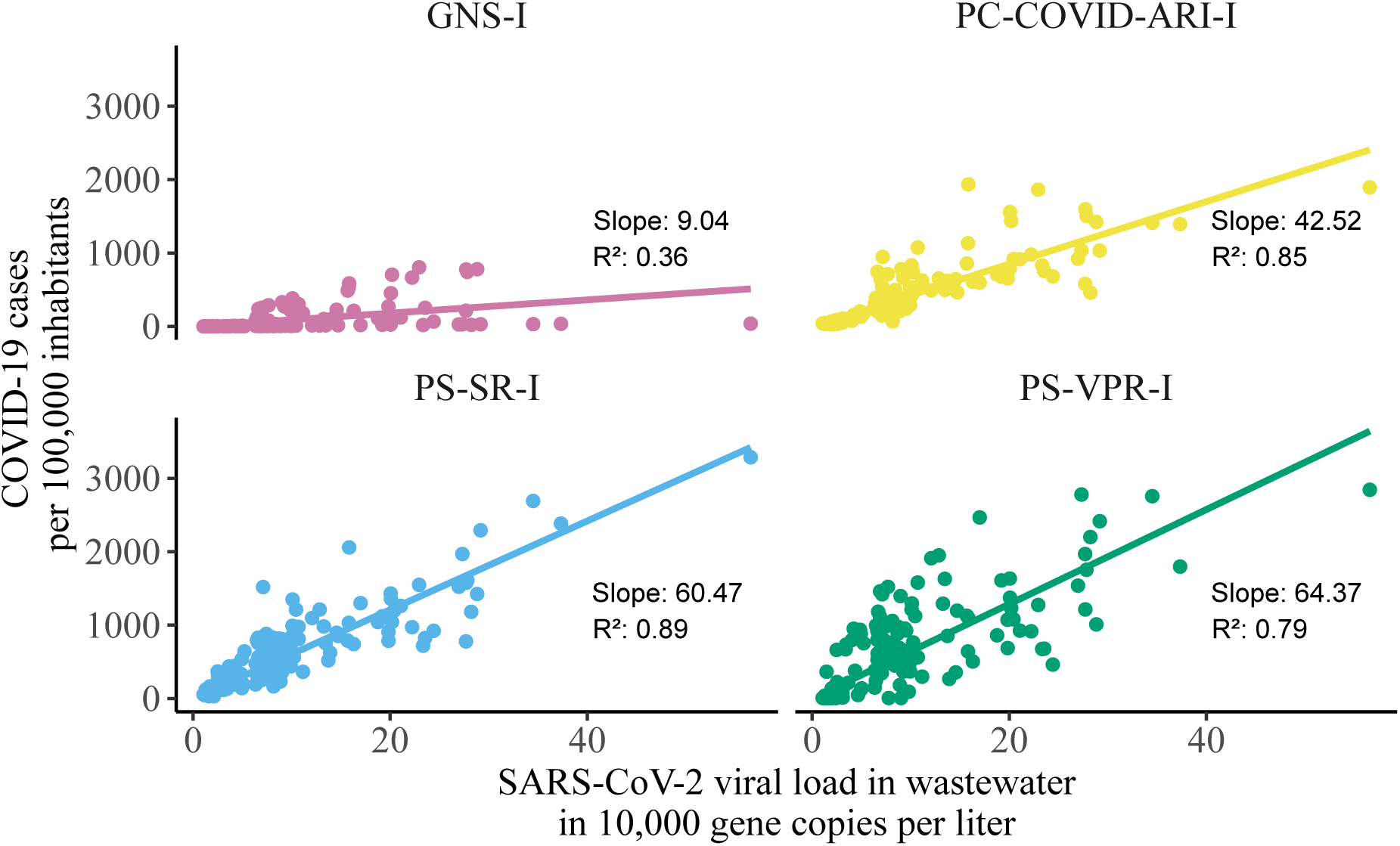
Relationship between wastewater viral load and reported incidence across four indicators. Each panel displays the linear association between weekly wastewater SARS-CoV-2 concentrations and corresponding incidence estimates, with regression lines and coefficients of determination. GNS-I = German Notification System of COVID-19, PC-COVID-ARI-I = Sentinel for Electronic Recording of Diagnoses of Acute Respiratory Infections, combined with Participatory System, PS-SR-I = Participatory System Self-Reported, PS-VPR-I = Participatory System Virological Positivity Rate.

### 3.3 Question 3: Do historical week-to-week trends in wastewater agree with historical week-to-week trends in reported incidence?

In Figure 7, the visual inspection of the time series plot of the changes from week-to-week shows that the wastewater viral load curve largely had a synchronous course compared with that of the week-to-week changes of PS-SR-I, PC-COVID-ARI-I, and GNS-I, indicating similar temporal dynamics. The PS-VPR-I (green) series also followed the general trend, but showed notable deviations in amplitude, especially in mid-2023 and early 2024.

**Figure 7:**
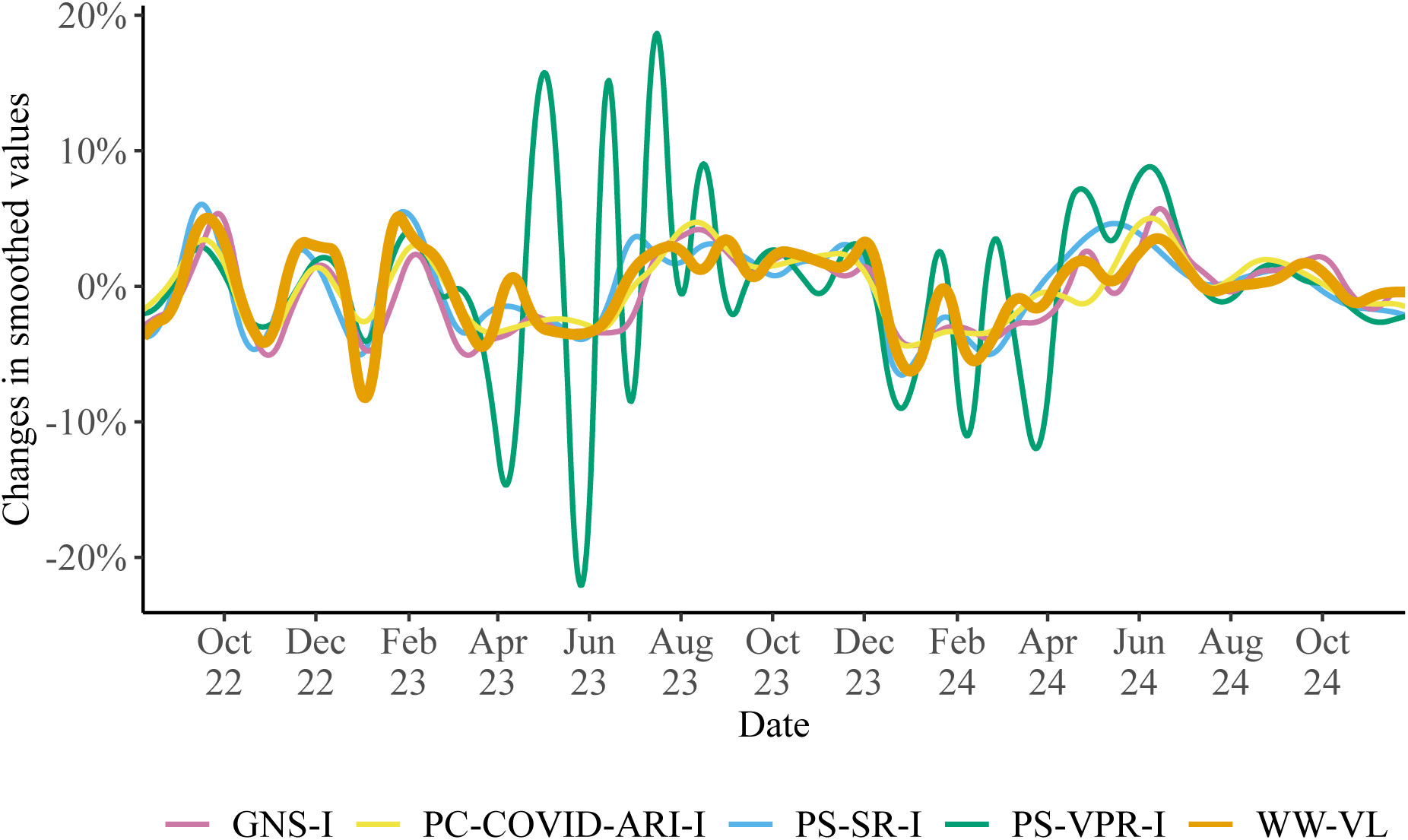
Week-to-week percentage changes in smoothed values of SARS-CoV-2 wastewater viral load and reported incidence indicators (2022-2024). GNS-I = German Notification System of COVID-19, PC-COVID-ARI-I = Sentinel for Electronic Recording of Diagnoses of Acute Respiratory Infections, combined with Participatory System, PS-SR-I = Participatory System Self-Reported, PS-VPR-I = Participatory System Virological Positivity Rate, WW-VL = Wastewater Viral Load.

These observations are supported by corresponding Pearson cross-correlation coefficients (Figure 8). No relevant time leads or lags were observed between the systems, indicating that changes in wastewater viral load and disease incidence, in particular as measured by PS-SR-I, GNS-I, and PC-COVID-ARI-I, occurred in a largely synchronous manner. The contemporaneous correlation coefficients were fairly high for these three systems (ranging from 0.82 to 0.88), while all cross-correlations were low for PS-VPR-I (0.37 for the contemporaneous correlation).

**Figure 8:**
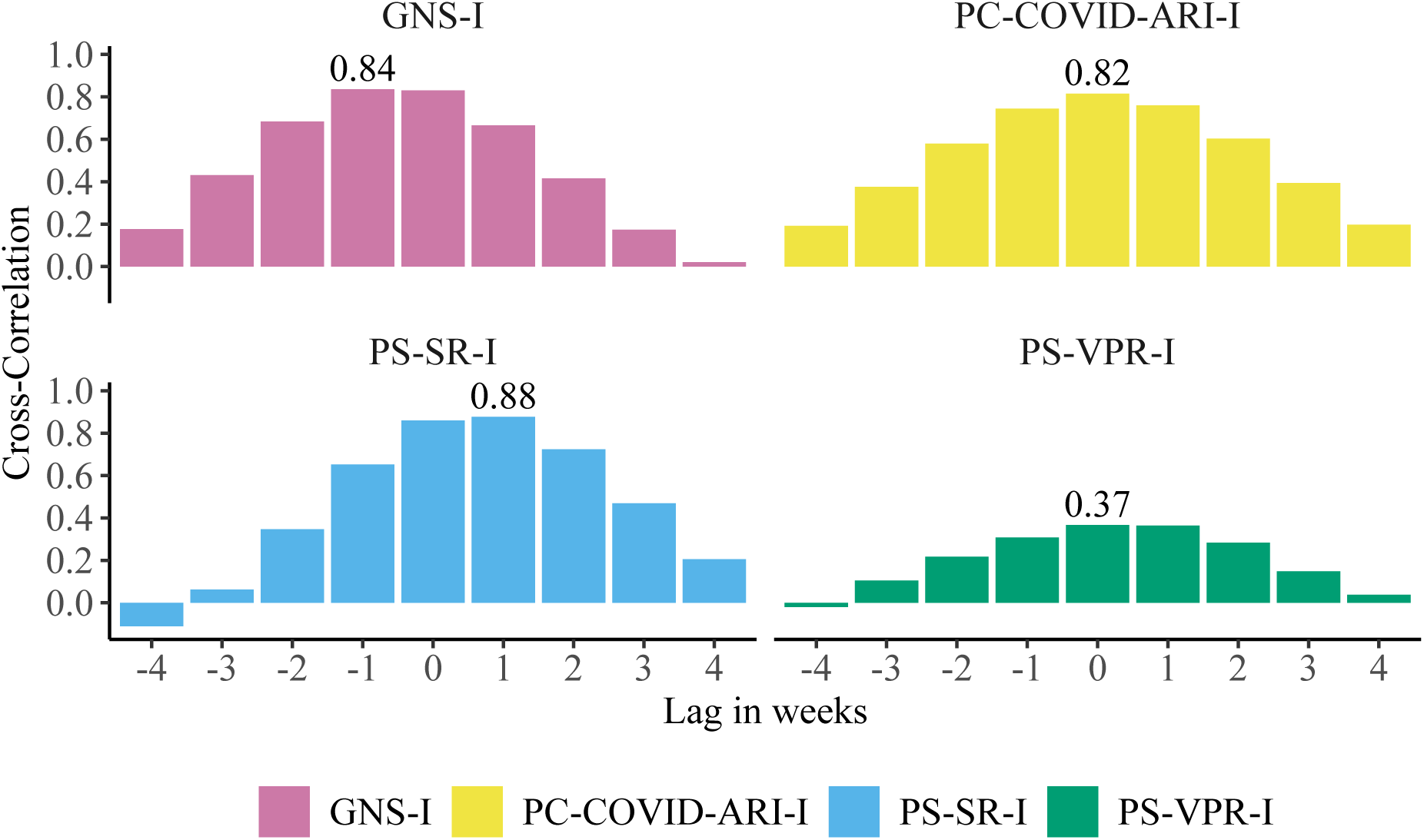
Cross-correlation between changes in smoothed wastewater viral load and four COVID-19 incidence indicators at varying lags. A correlation for a lag of two, e.g., indicates the correlation between changes in smoothed incidence and changes in smoothed wastewater viral load that were shifted two weeks back in time. GNS-I = German Notification System of COVID-19, PC-COVID-ARI-I = Sentinel for Electronic Recording of Diagnoses of Acute Respiratory Infections, combined with Participatory System, PS-SR-I = Participatory System Self-Reported, PS-VPR-I = Participatory System Virological Positivity Rate.

After categorizing changes into week-to-week trends (Increase, Decrease, Stable), we see that trends in wastewater agree in at least three out of the four case-based surveillance systems in 74% of the weeks (Table 1). In this retrospective consideration, 90% (80%) of the increases (decreases) in at least three out of the four case-based surveillance systems were also observed in wastewater viral load (sensitivity), while 79% (95%) of the increases (decreases) in wastewater were reflected by the same trend in at least three out of the four case-based surveillance systems (positive predictive value). Non-agreement occurred mostly close to plateaus in the wastewater curve (Figure 9).

**Figure 9:**
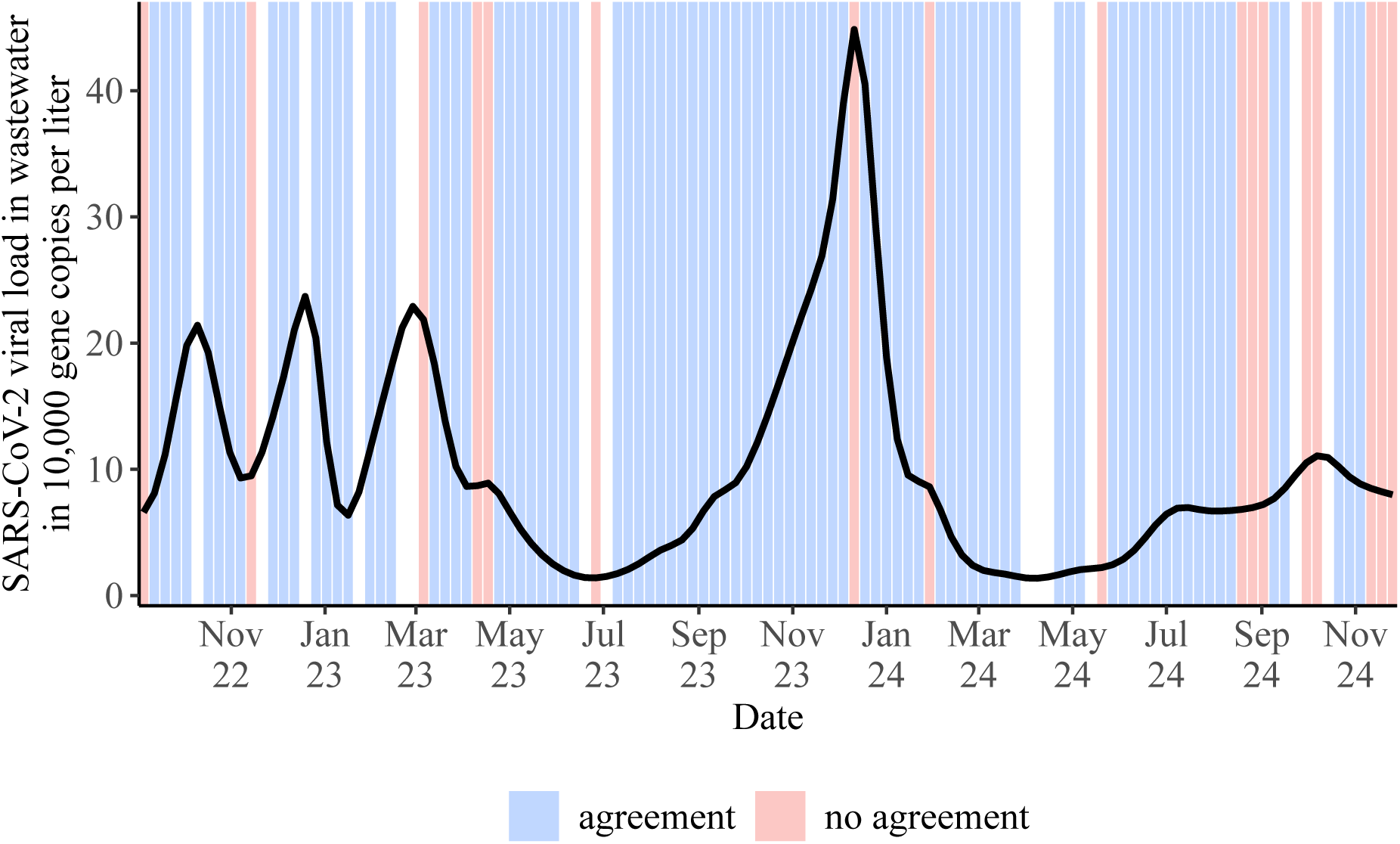
Agreement between week-to-week trends in wastewater viral load and trends in at least three of the four case-based surveillance systems (retrospective consideration) over time. Shown are the smoothed wastewater viral loads and weeks exhibiting agreement (blue) and no agreement (red). Blank spaces indicate weeks without clear trend indicated by the case-based surveillance systems (category ”Unclear”).

**Table 1:**
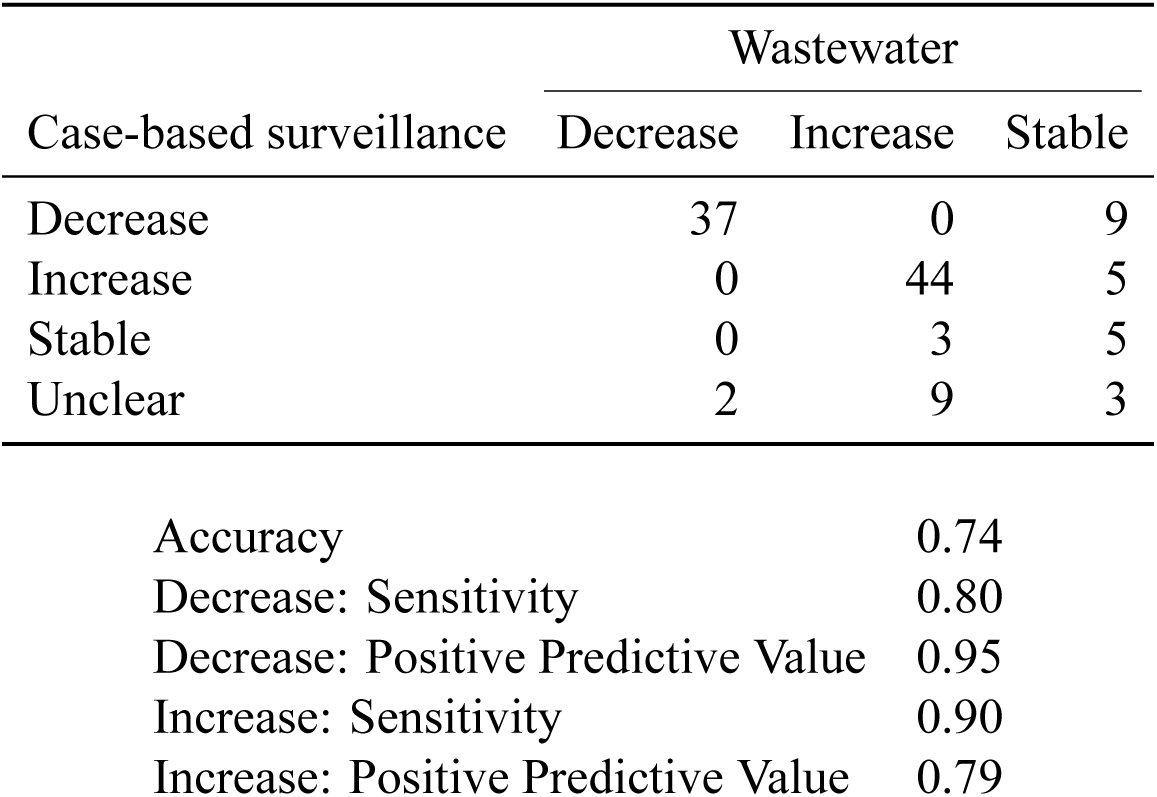
Contingency table between week-to-week changes in wastewater viral load and week-to-week changes in at least three of the four case-based surveillance systems (retrospective consideration) between week 36/2022 and week 48/2024. Corresponding summary statistics are reported below. ”Increase”/”Decrease” = change of more than 5% compared to previous week, ”Stable” = change of less than 5%, ”Unclear” = no agreement among more than two case-based surveillance systems.

| Case-based surveillance | Wastewater |  |  |
| --- | --- | --- | --- |
|  | Decrease | Increase | Stable |
| Decrease | 37 | 0 | 9 |
| Increase | 0 | 44 | 5 |
| Stable | 0 | 3 | 5 |
| Unclear | 2 | 9 | 3 |

|  |  |
| --- | --- |
| Accuracy | 0.74 |
| Decrease: Sensitivity | 0.80 |
| Decrease: Positive Predictive Value | 0.95 |
| Increase: Sensitivity | 0.90 |
| Increase: Positive Predictive Value | 0.79 |

### 3.4 Question 4: Can wastewater data be used to predict disease incidence trends for the current week?

The mean inner loop cross-validation overall accuracy was highest for the LASSO (69%, Table A3). Considering the test sets (the hold-out-sets from the outer loop), a similar performance was observed (Table 2): the overall accuracy was 68%, implying that if a certain trend (Increase, Decrease, Unclear) was predicted by the LASSO using wastewater data, the prediction was reflected by the case-based surveillance systems with 68% probability. In particular, 80% (87%) of the increases (decreases) in at least three out of the four case-based surveillance systems were correctly predicted by the model (sensitivity), while 68% (67%) of the predicted increases (decreases) were correctly mirrored by the same trend in at least three out of the four case-based surveillance systems (positive predictive value). None of the ”Other” trends (grouping the categories ”Stable” and ”Unclear”) in the case-based surveillance systems were correctly predicted by the LASSO.

**Table 2:**
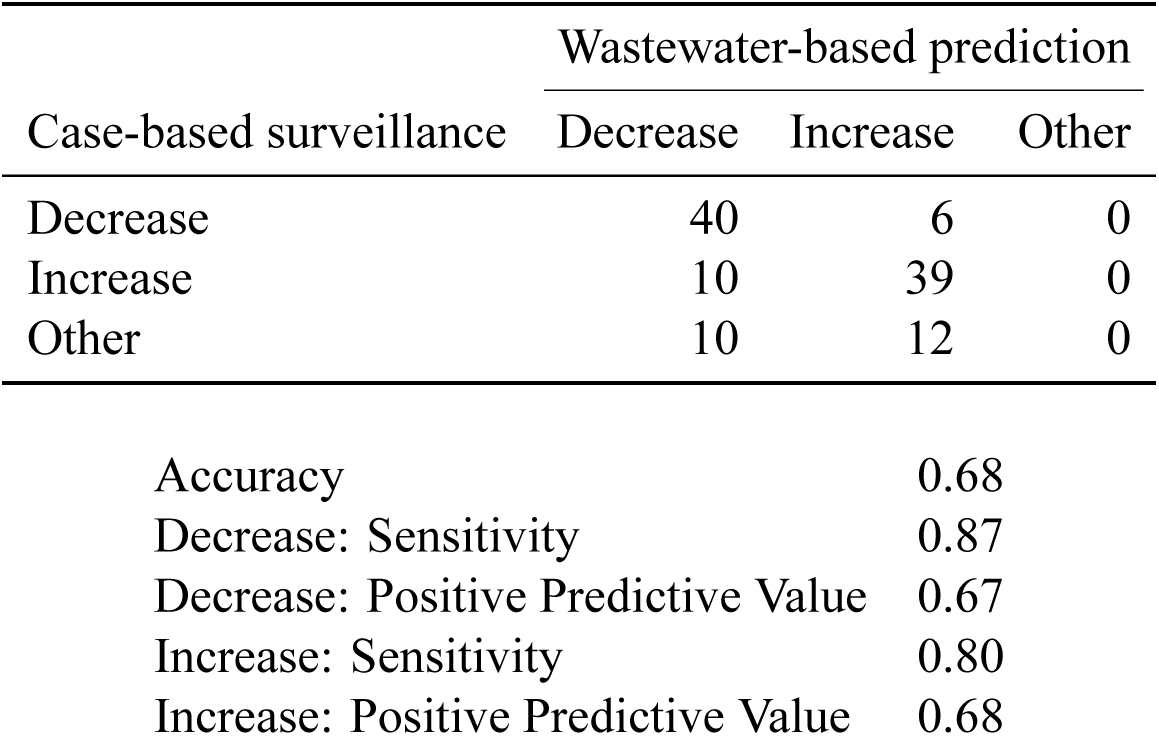
Contingency table comparing LASSO predictions and actual week-to-week changes in at least three of the four case-based surveillance systems (current-week prediction) between week 36/2022 and week 48/2024. Corresponding summary statistics are reported below. ”Increase”/”Decrease” = change of more than 5% compared to previous week, ”Other” = Change of less than 5% or no agreement among more than two case-based surveillance systems.

| Case-based surveillance | Wastewater-based prediction |  |  |
| --- | --- | --- | --- |
|  | Decrease | Increase | Other |
| Decrease | 40 | 6 | 0 |
| Increase | 10 | 39 | 0 |
| Other | 10 | 12 | 0 |

|  |  |
| --- | --- |
| Accuracy | 0.68 |
| Decrease: Sensitivity | 0.87 |
| Decrease: Positive Predictive Value | 0.67 |
| Increase: Sensitivity | 0.80 |
| Increase: Positive Predictive Value | 0.68 |

## 4. Discussion

This study assessed the capability of viral load data measured in wastewater to track, estimate, and predict COVID-19 incidence and its dynamics. Results were evaluated and compared with data from four case-based surveillance systems. Our analysis addressed several key questions, including the responsiveness of wastewater data to changes in disease incidence, its retrospective utility in estimating disease incidence, its agreement with historical trends, and its potential to predict current week-to-week trends in disease incidence. The findings show that the viral load in wastewater closely agrees with disease incidence, with no relevant lead or delays, including periods of rapid increases and even periods of rapid decreases in incidence. Wastewater viral load can be retrospectively translated into disease incidence on average, but is not adequate for predicting disease incidence in real-time. Historical week-to-week trends are largely well reflected in wastewater data. Wastewater data can also be used, to a moderate degree, to predict trends.

### 4.1 Question 1: How well does wastewater viral load correspond to disease incidence overall and particularly in terms of timeliness, increases, and decreases?

The first objective of this study is to determine how well wastewater data agree with rises and falls in disease incidence. GNS-I diverges increasingly from all other systems by showing progressively lower case incidence, likely due to under-ascertainment due to less frequent testing especially since 2023 [Loenenbach et al., 2024]. However, wastewater data agree well with the other three case-based surveillance data (PS-SR-I, PS-VPR-I, and PC-COVID-ARI-I), indicating that wastewater viral loads generally mirror changes in population-level disease dynamics, especially during major surges or declines in incidence. This is consistent with findings from the systematic review and meta-analysis study by Li et al. [2023], which showed that viral loads in wastewater strongly correlate with reported incidence.

While some studies have also reported good agreement between wastewater data and prevalence data [Mohring et al., 2024, Layton et al., 2022], Li et al. [2023] concluded that the correlation between normalized viral RNA in wastewater and new (incident) cases is stronger than with active (prevalent) cases. In this study, we found no meaningful differences when comparing wastewater data with either incidence data or prevalence-like estimates based on shedding-derived curves. This may reflect the limited ability of current systems to detect subtle differences between incidence and prevalence, which is plausible given the relatively short shedding period of SARS-CoV-2 (Figure 3).

The correlation coefficients in our analysis (0.73-0.87) for PS-SR-I, PS-VPR-I, and PC-COVID-ARI-I place our findings at the upper end of the range of the studies reported by Li et al. [2023], indicating very good agreement. In contrast, GNS-I did not correlate as well with wastewater data as the other systems, likely due to changes in PCR testing patterns during the study period [Loenenbach et al., 2024].

The different case-based surveillance systems presented in this study vary in their dependencies on external factors (Table A1), including self-testing behavior (PS-SR-I), testing behavior by health care providers (PC-COVID-ARI-I, GNS-I), and health care-seeking behavior (GNS-I, PS-VPR-I, PC-COVID-ARI-I), all of which may change over time. However, the consistency of trends across several independent systems reinforces the validity of their results. The high level of agreement indicates that, during the study period, all analyzed case-based surveillance system, except GNS-I, closely reflected the unknown true incidence and were suitable for detecting disease dynamics.

Time lags between wastewater and clinical data may depend on factors such as time, region, population, access to testing, and the type of clinical indicator [Chen et al., 2024], see also Table A1. Systems or indicators that capture incidence or events that follow with a certain time span after the time of infection are generally expected to show a lag in cross-correlation. For example, Hopkins et al. [2023] reported that while wastewater viral load and syndromic surveillance data were consistent in timing, they had a lead time compared to changes in COVID-19-related general and intensive care bed use rates by one to two weeks.

We observed no relevant time lags overall, though several points merit consideration. First, the time lag analysis is retrospective in nature and does not account for reporting delays in any of the systems, such as the time between wastewater sampling and data availability at Robert Koch Institute. Second, any time lag is most important during periods of rapidly rising incidence. We observed overall synchronicity, although the slope of the viral load curve was sometimes steeper than that of the case-based systems and sometimes not. Thus, based on our data, wastewater surveillance demonstrates comparable timeliness to case-based surveillance in monitoring SARS-CoV-2/COVID-19. Third, behavior during declining incidence is of particular interest, because these periods would most clearly reveal whether wastewater data lag due to prolonged viral shedding [Gerrity et al., 2020]. We tested this point by comparing PS-SR-I with a hypothetical prevalence curve derived from PS-SR-I and found a negligible shift, even during periods of rapid decline such as during January 2024 (Figure 4). Overall, our finding confirms that wastewater data correlate well with both incidence and prevalence data [Mohring et al., 2024, Li et al., 2023, Róka et al., 2021] but does not imply that WBS can be used as an early warning system for SARS-CoV-2.

### 4.2 Question 2: Can wastewater data be retrospectively translated into an estimate of disease incidence?

We demonstrated a strong linear correlation between smoothed wastewater SARS-CoV-2 concentrations and smoothed incidence estimates from PS-SR-I, PS-VPR-I, and PC-COVID-ARI-I (Figure 6). The weakest correlation was observed with the notification-based indicator GNS-I (R² = 0.36), likely due to reduced testing frequency and consequently increasing under-ascertainment [Loenenbach et al., 2024]. During the first two years of the pandemic, GNS-I has reflected incidence much accurately [Loenenbach et al., 2024]. This highlights the influence of behavior- and policy-related pandemic, phase–specific factors on surveillance sensitivity, from which wastewater surveillance is ”immune” to. The high level of agreement between wastewater data and the two main participatory system-based indicators PS-SR-I and PS-VPR-I suggests that systems closely reflect the actual magnitude of disease at the population level. Both indicators measure disease dynamic independently. PS-VPR-I also incorporates inputs from virological surveillance. The two systems produced the highest and nearly identical translation factors.

However, even for our most stable indicator, PS-SR-I, the translation factor varied by up to a factor of five over time (Figure 5). This week-to-week fluctuation suggests considerable uncertainty in real-time estimates. Sources of this variation likely include fluctuations in both wastewater and case-based surveillance data. For instance, changes in testing rates (GNS-I, PS-SR-I), sample size in virological surveillance (PS-VPR-I), healthcare-seeking behavior (PC-COVID-ARI-I), and variant dynamics may all contribute [Yang et al., 2025, Huisman et al., 2022, Bar-Or et al., 2022]. Wastewater viral load may also be affected by variant-specific shedding patterns [Yang et al., 2025]. For these reasons, it might not be advisable to replace case-based data by wastewater-based incidence estimates.

More advanced models that incorporate additional information might help explain week-to-week fluctuations in the translation factor and reduce uncertainty of derived incidence estimates. Such information could include testing rates [McManus et al., 2023], spatial data [Cuadros et al., 2024], or mobility data [Schenk et al., 2024]. More generally, previous work has shown that wastewater data can provide moderate benefit in estimating incidence or prevalence, particularly when clinical surveillance data are sparse or absent [Li et al., 2024, Klaassen et al., 2024].

### 4.3 Question 3: Do historical week-to-week trends in wastewater agree with historical week-to-week trends in reported incidence?

Our findings show that week-to-week changes in SARS-CoV-2 viral load in wastewater closely and retrospectively agree with week-to-week trends from three of the four case-based surveillance systems (Figure 7): PS-SR-I, PC-COVID-ARI-I, and GNS-I. This suggests that wastewater can reliably reflect short-term shifts in community infection dynamics. The agreement was less evident for PS-VPR-I, which displayed more erratic fluctuations, particularly during periods of low case numbers. This is likely because PS-VPR-I is based on the virological positivity rates, which can vary considerably when weekly sample sizes are smaller during low incidence periods. Notably, although GNS-I has lost its ability to reflect the number of cases in the population over time, its week-to-week percent changes closely agree with these of the other systems (except PS-VPR-I) throughout the whole observation period.

These correlations are further supported by Pearson cross-correlation analyses of the week-to-week changes (Figure 8), where we identified no relevant time lag. This indicates that increases and decreases in disease activity were captured nearly simultaneously by wastewater and case-based surveillance systems, albeit with some variability in timing. A similar relationship between week-to-week changes in wastewater data and case-based indicators was observed in a prior study [Amato et al., 2023], which analyzed periods of increasing and decreasing incidence separately. That study reported that correlations peaked at a 1–2 week lead time for wastewater data during periods of increasing incidence and approximately zero lag during periods of decline, further supporting the responsiveness of wastewater surveillance to dynamic shifts in infection trends.

The trend-based categorization added interpretability beyond correlation coefficients. In 74% of weeks, wastewater trends matched the direction of change in at least three out of the four case-based systems. This high level of agreement in (historical) trends (sensitivity of 90% for increases, 80% for decreases; positive predictive value of 79% for increases, 95% for decreases) indicates that wastewater can serve as a robust indicator of both rising and declining trends in disease activity. Most mismatches in changes between wastewater and the cased-based surveillance systems originate from the more frequent occurrence of ”Stable” classification in wastewater viral load and in the ”Unclear” category in the case-based surveillance systems, which has no equivalent in wastewater viral load. These mismatches primarily occurred around peaks or troughs and at the end of the study period, when increases and decreases in wastewater viral load were less pronounced (Figure 9).

### 4.4 Question 4: Can wastewater data be used to predict disease incidence trends for the current week?

We also emulated the use of real-time wastewater data to predict the trend of disease incidence in the current week. Our results showed that an (unobserved) increase (decrease) in the current week in at least three of the four case-based surveillance systems would be predicted by the selected machine learning model with 80% (87%) probability (Table 2). Moreover, if the model predicted an increase (decrease) in incidence, this would be reflected in at least three of the four case-based surveillance systems with 68% (67%) probability. These positive predictive values are comparatively low since none of the trends classified as ”Other” could be correctly predicted. The reason for this is that ”Other” comprises two different small categories, with the larger one (”Unclear”) indicating disagreement between the case-based surveillance system. This issue could only be avoided if agreement between the case-based surveillance systems was higher.

Overall, while we found that retrospective translation from wastewater viral load to SARS-CoV-2 disease incidence is associated with large uncertainty on a weekly basis (Question 2), the results of our model predicting changes in disease incidence appear more convincing. Such a model may be useful if case-based systems have lost validity or are no longer maintained. However, wastewater-based trend prediction models require calibration. To this end, either robust case-based systems, as applied in this study, or a temporary high-quality cohort as in Mohring et al. [2024] may serve as a reference. As noted in the discussion of Question 2, additional information such as testing rates, spatial or mobility data could improve the predictive power of these models.

From a broader perspective, wastewater can only add certain aspects to the overall necessary epidemiological picture. Its strengths include low-cost quantitative information on the infection burden in the population, on the occurrence of infections at fine spatial levels, detection of trends, short-term trend prediction, and the description of the proportion and dynamics of variants. Based on our experience, the translation of wastewater viral load to incidence is mainly possible only in retrospect. The collection of other information about affected individuals, such as age, gender, symptoms, or disease severity, remains indispensable and must continue to rely on traditional case-based systems.

### 4.5 Limitations

One limitation concerns the gradual expansion of the WBS system during the study period. This entailed a continuous augmentation of participating wastewater treatment plants, increasing from 36 to 161 over the study period. Despite this expansion, deviations from the smoothed curve remained stable over time, indicating that national averages are not heavily influenced by the addition of new plants (Figure 1).

In all analyses, we did not adjust for reporting delay, which affected all systems to some extent. While this was not critical for retrospective considerations, real-time predictions, as addressed in Question 4, should be interpreted with caution, as the wastewater data used here do not fully reflect what would have been available in real time. In practice, it is essential to ensure that contributing wastewater treatment plants provide their data promptly and consistently over time.

Finally, the use of routine surveillance data introduces inherent limitations that must be considered (Table A1). Although these data do not match the quality compared to data if they had been collected in the framework of, e.g. repeated prevalence surveys, such as those conducted by Mohring et al. [2024], the combined use of four different systems likely provided a robust estimate of true incidence.

## 5. Conclusion

By comparing wastewater viral load data to four distinct case-based surveillance systems in Germany from mid-2022 to the end of 2024, we found that wastewater data have closely mirrored changes in disease incidence at the population level, with virtually no time lag. On average, wastewater data and disease incidence exhibited strong correlations for three of the four case-based surveillance systems investigated. However, translating wastewater viral load into precise real-time incidence was associated with relatively large week-to-week variation estimates remains challenging due to temporal fluctuations and system-level variability which might attributable to variability within the case-based surveillance systems, the wastewater surveillance, or both. Due to this variation, we do not recommend using wastewater data for real-time incidence estimation, or at least not in isolation and only with considerable caution. While wastewater data largely aligned retrospectively with the week-to-week trends (changes) observed in at least three of the four case-based surveillance systems, machine learning models showed moderate success in predicting current-week trends. To this end, models using wastewater data might be useful to cautiously predict the course of the incidence for the current week, particularly when case-based surveillance systems are unreliable or not available, but developing more nuanced models including additional relevant variables is recommended. Overall, this study demonstrates the promise of wastewater-based surveillance as a valuable, complementary tool for monitoring COVID-19 dynamics and encourages the use of wastewater monitoring for additional pathogens.

## Acknowledgments

We appreciate the support of the Federal Ministry of Health, which has funded the wastewater surveillance project, *Abwassermonitoring für die epidemiologische Lagebewertung* (AMELAG), through the end of 2025. We sincerely appreciate the support and dedication of the entire AMELAG team. We would like to recognize the invaluable contributions of Ulrike Braun, Marcus Lukas, and all participating wastewater treatment plants, laboratories, academic institutions, and state authorities associated with the AMELAG initiative.

Our thanks go to the dedicated staff of the acute respiratory infections team at the Respiratory Infections Unit of the Robert Koch Institute (including Walter Haas, Julia Schilling, Kristin Tolksdorf, Simon Krupka, and Juliane Wunderlich) as well as to Michaela Diercke and Jeremias Weber from the Surveillance and Electronic Reporting and Information System Unit, for their valuable comments and support. We are also grateful to Ralf Dürrwald and the personnel at the National Influenza Centre.

Finally, We extend our sincere gratitude to all individuals who participated in GrippeWeb, the primary care sentinel physicians involved in the syndromic surveillance SEED/ARE and the virologic surveillance of the physician sentinel (conducted by the National Reference Center for Influenza), as well as the local health authorities who collected information on cases of COVID-19 and provided notification data.

## Abbrevations

ARI: Acute Respiratory Infections
GNS-I: German Notification System Incidence
ILI: Influenza-like Illness
LASSO: Least Absolute Shrinkage and Selection Operator
PC-COVID-ARI-I: Primary Care-COVID-ARI-I, Sentinel for Electronic Recording of Diagnoses of Acute Respiratory Infections, combined with Participatory System
PR: Positivity Rates
PS-SR-I: Participatory System Self-Reported
PS-VPR-I: Participatory System Virological Positivity Rate
R²: Coefficients of Determination
WBS: Wastewater-Based Surveillance
WWTPs: Wastewater Treatment Plants
WW-VL: Wastewater Viral Load

## Data Availability Statement

Data and software code for replicating the analyses are available on https://github.com/peterpuetz2020/WBS_for_COVID19_monitoring.

## Ethical Standards

The research meets all ethical guidelines, including adherence to the legal requirements of the study country.

## Author Contributions

Conceptualization: S.A., U.B., P.P., J.S.; Methodology: S.A., U.B., P.P., J.S.; Data acquisition: S.A., U.B., T.G., AS.L., P.P., A.S., J.S.; Data curation: P.P.; Data visualisation: P.P.; Writing – review and editing: S.A., U.B., T.G., AS.L., P.P., A.S., J.S.; Writing original draft: S.A., U.B., P.P. J.S. All authors approved the final submitted draft.

## Appendix

**Table A1:** Description and comparison of the four cased-based surveillance systems considered in this study.

|  | German<br>Notification System<br>of COVID-19<br>(GNS-I) | Participatory<br>System<br>Self-Reported<br>(PS-SR-I) | Participatory System<br>Virological<br>Positivity Rate<br>(PS-VPR-I) | Sentinel for<br>Electronic<br>Recording of<br>Diagnoses of<br>Acute<br>Respiratory<br>Infections,<br>combined with<br>Participatory<br>System<br>(PC-COVID-ARI-I) |
| --- | --- | --- | --- | --- |
| Captured<br>indicator | SARS-CoV-2<br>infection incidence | COVID-19<br>incidence | COVID-19<br>incidence | COVID-19<br>incidence |

Table A1: Description and comparison of the four case-based surveillance systems considered in this study.
|  | German Notification System of COVID-19 (GNS-I) | Participatory System Self-Reported (PS-SR-I) | Participatory System Virological Positivity Rate (PS-VPR-I) | Sentinel for Electronic Recording of Diagnoses of Acute Respiratory Infections, combined with Participatory System (PC-COVID-ARI-I) |
| --- | --- | --- | --- | --- |
| How does it work? | - Mandatory reporting System.<br>- Physicians and other entities report suspected, confirmed, and deceased COVID-19 cases.<br>- Laboratories report lab confirmations of acute infections.<br>- Local public health agencies combine lab, physician and other notifications and confirm the cases.<br>- Cases are counted if they fulfill the case-definition by the Robert Koch Institute [Robert Koch Institut, 2023]. | - Based solely on GrippeWeb.<br>- Participants self-report positive COVID-19 test results (from physicians or self-testing).<br>- COVID-19 incidence is estimated by dividing positive tests by total weekly report.<br>- Available since summer 2022. | - Based on two systems:<br>(1) Virological surveillance in the primary care system, where sentinel physicians collect swabs from ARI patients and send them to the National Reference Laboratory for Influenza at the RKI.<br>(2) GrippeWeb, a web-based system where every citizen can register and report weekly on the presence or absence of ARI. Two syndromes are calculated: ARI and Influenza-like Illness (ILI).<br>- COVID-19 Incidence is calculated by combining ILI and non-ILI rates attributed to COVID-19 [Buchholz et al., 2023] | - Sentinel system coordinated by RKI.<br>- Physicians enter lab-confirmed COVID-19 cases into their electronic information systems using ICD Code U07.<br>- Approximately 200–250 physicians report lab-confirmed COVID-19 cases weekly centrally to the RKI.<br>- Incidence is estimated by adjusting the proportion of ill persons who seek care (based on data from GrippeWeb). |

Table A1: Description and comparison of the four case-based surveillance systems considered in this study.
|  | German Notification System of COVID-19 (GNS-I) | Participatory System Self-Reported (PS-SR-I) | Participatory System Virological Positivity Rate (PS-VPR-I) | Sentinel for Electronic Recording of Diagnoses of Acute Respiratory Infections, combined with Participatory System (PC-COVID-ARI-I) |
| --- | --- | --- | --- | --- |
| Week of analysis refers to | - Reporting date. | - Week of illness. | - Week of illness. | - Week of consultation, covering illnesses from the previous two weeks. |
| Strengths | - Mandatory reporting.<br>- Comprehensive.<br>- High specificity (PCR-confirmed only).<br>- Timely reporting (days).<br>- Well-established system. | - Stable estimate since summer 2022.<br>- Independent of physicians' test-ordering and consultation behavior.<br>- Quite specific due to use of antigen tests.<br>- Sensitivity varies by test.<br>- Fast (approximately one-week delay from symptom onset). | - Estimated incidence is independent of testing and healthcare seeking behavior.<br>- Virological testing occurs after swab collection.<br>- High specificity due to laboratory testing. | - Weekly actively report by sentinel physicians. |

|  | German Notification System of COVID-19 (GNS-I) | Participatory System Self-Reported (PS-SR-I) | Participatory System Virological Positivity Rate (PS-VPR-I) | Sentinel for Electronic Recording of Diagnoses of Acute Respiratory Infections, combined with Participatory System (PC-COVID-ARI-I) |
| --- | --- | --- | --- | --- |
| Threats | - Largely passive system.<br>- Infected individuals have to undergo PCR testing, but testing behavior changed massively during the study period due to changes in reimbursement, setup and closure of testing facilities, self-testing, and shifting rules.<br>- Depends on overwhelmed local public health agencies during peaks. | - Largely passive system.<br>- Depends on participants purchasing and using SARS-CoV-2 tests themselves.<br>- Motivation to test may decrease post-pandemic.<br>- Selection bias: Participants are likely to be health-conscious individuals.<br>- Relies on self-reported data. | - Largely passive system.<br>- Positivity rate might be affected if patients with positive self-tests avoid consulting physicians.<br>- Relies on self-reported data. | - Largely passive system.<br>- Dependent on patients consulting a physician.<br>- Mild cases may be missed (though consultation proportion adjusts for that).<br>- Risk that patients with a positive self-test do not visit a physician.<br>- Depends on physicians to code and test correctly<br>- Depends on whether physicians order a test.<br>- Depends on the diagnosis-depth (e.g., stopping at ARI or not testing). |

Table A1: Description and comparison of the four cases-based surveillance systems considered in this study.
|  | German Notification System of COVID-19 (GNS-I) | Participatory System Self-Reported (PS-SR-I) | Participatory System Virological Positivity Rate (PS-VPR-I) | Sentinel for Electronic Recording of Diagnoses of Acute Respiratory Infections, combined with Participatory System (PC-COVID-ARI-I) |
| --- | --- | --- | --- | --- |
| Development since the beginning of the pandemic | - After sufficient test capacity became available (around autumn 2020), most COVID-19 cases were likely tested until end of 2021.<br>- From spring 2022, a growing testing gap due to reimbursement and regulation changes<br>- Since 2023, reported incidence has been likely underestimated and unrealistic. | - Relatively stable since data collection began in summer 2022. | - Relatively stable due to independence from test reimbursement, testing rules, and other testing practices. | - Since end of 2022, reimbursement for physician testing was reduced. |

**Table A2:** Comparison of Peak SARS-CoV-2 Viral Load Kinetics Over Time and Variants: Insights from Three Studies. SO = symptom onset. Note: Arts et al. [2023] used dry weight, while Wannigama et al. [2024] and Wölfel et al. [2020] used wet weight.

| Study name | No. of patients | Variants covered | Time of study | Peak Viral Load (Day after SO) |
| --- | --- | --- | --- | --- |
| Wannigama et al. [2024] | 113 | Omicron BA.2.86, JN.1, others | Aug 2022 – Dec 2023 | Days 5–10 |
| Arts et al. [2023] | 48 | Ancestral B.1.2, Alpha (B.1.1.7) | Dec 2020 – Mar 2021 | Days 3–4 |
| Wölfel et al. [2020] | 9 | Ancestral strain | Jan 2020 – Mar 2021 | Days 5–10 |

**Table A3:** Summary of inner loop cross-validation accuracies for the different Machine Learning classifiers.

| Classifier | Min. | 1st Qu. | Median | Mean | 3rd Qu. | Max. |
| --- | --- | --- | --- | --- | --- | --- |
| LASSO | 0.63 | 0.67 | 0.69 | 0.69 | 0.71 | 0.75 |
| Support Vector Machine | 0.59 | 0.62 | 0.64 | 0.64 | 0.65 | 0.70 |
| Random Forest | 0.61 | 0.64 | 0.68 | 0.67 | 0.70 | 0.73 |

**Figure A1:**
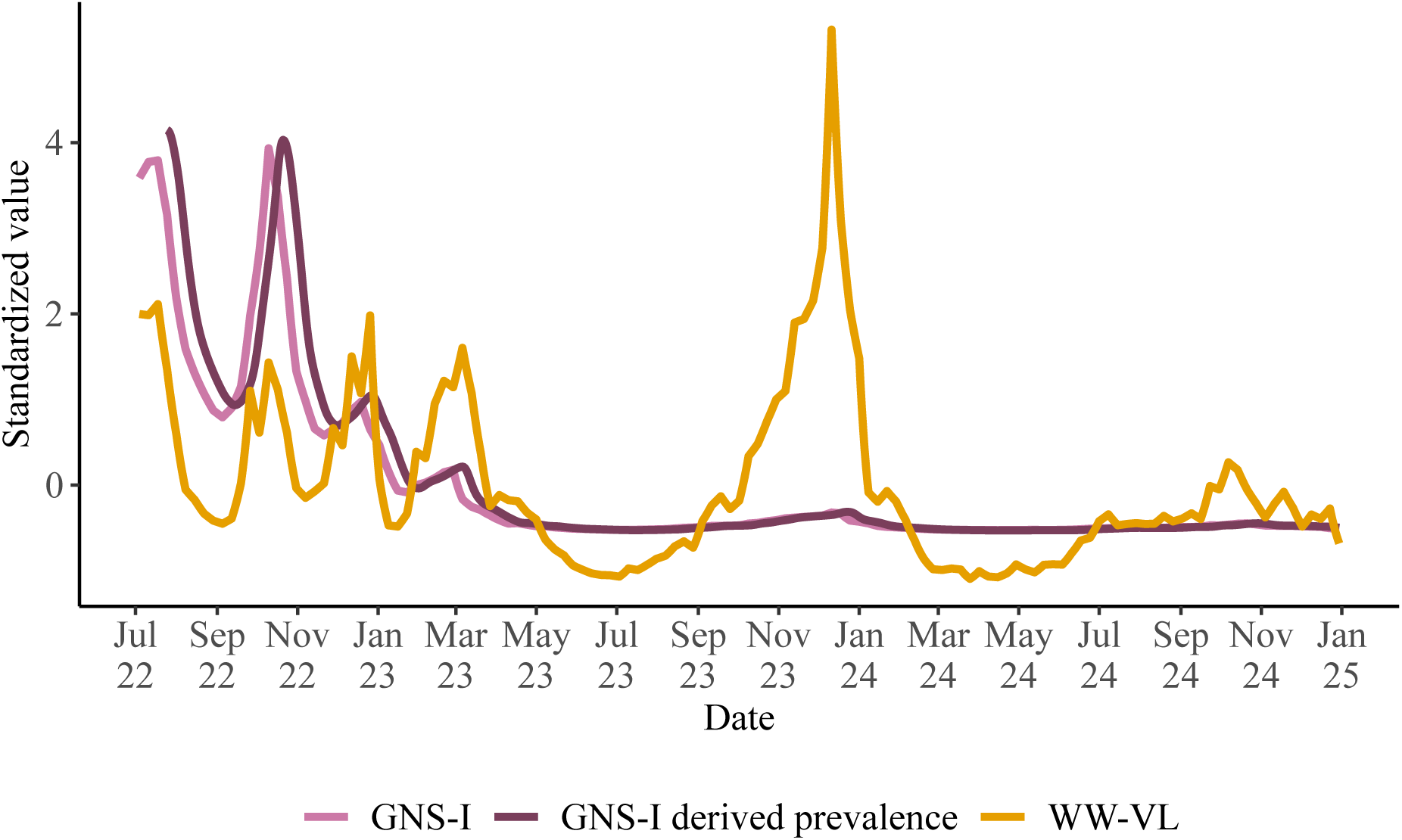
Hypothetical prevalence for GNS-I. GNS-I incidences were multiplied with the fecal shedding distribution shown in Figure 3 to derive a hypothetical prevalence curve. Shown are standardized values for the resulting hypothetical prevalence, the original incidence for GNS-I and the viral load in wastewater. GNS-I = German Notification System of COVID-19, WW-VL = Wastewater Viral Load.

**Figure A2:**
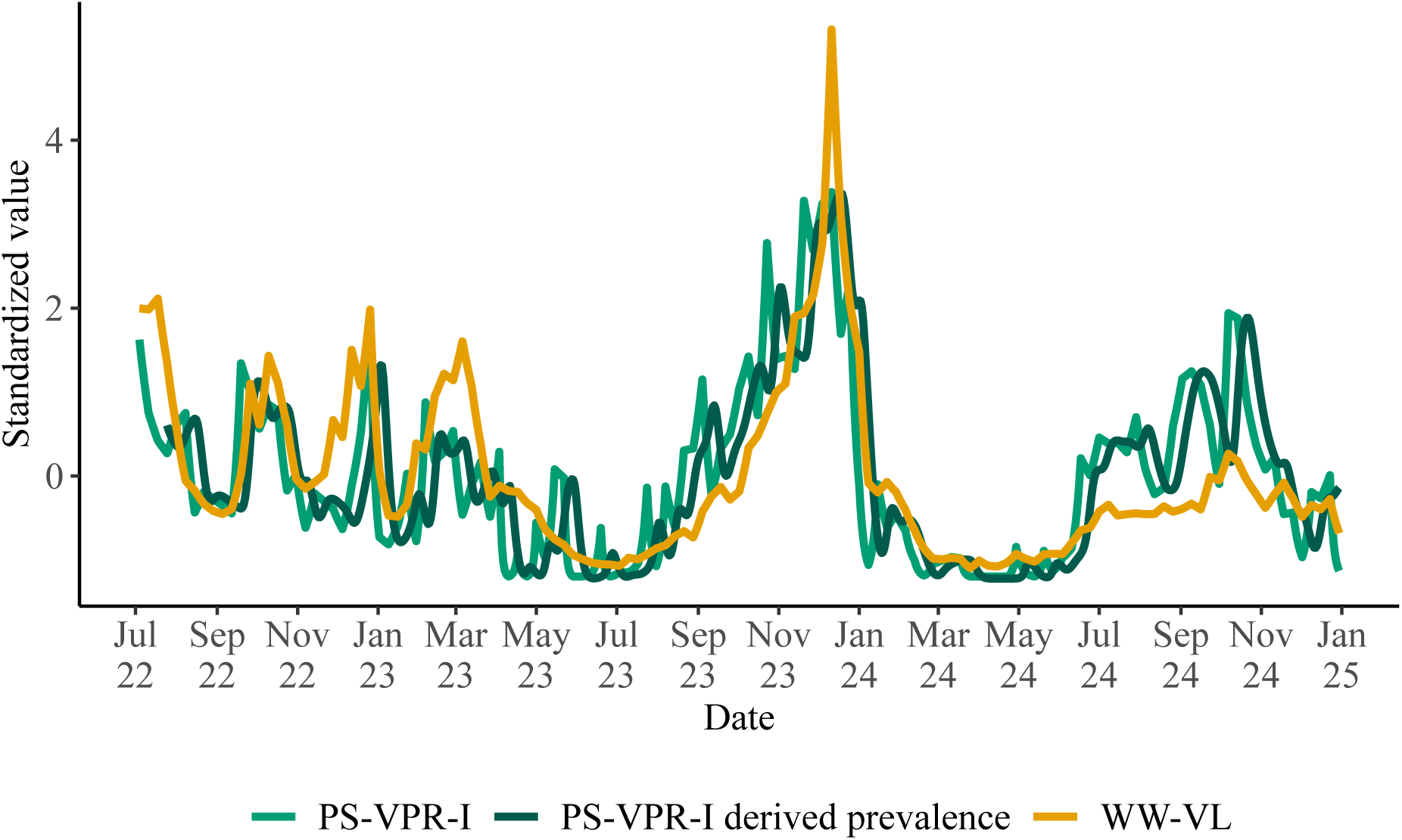
Hypothetical prevalence for PS-VPR-I. PS-VPR-I incidences were multiplied with the fecal shedding distribution shown in Figure 3 to derive a hypothetical prevalence curve. Shown are standardized values for the resulting hypothetical prevalence, the original incidence for PS-VPR-I and the viral load in wastewater. PS-VPR-I = Participatory System Virological Positivity Rate, WW-VL = Wastewater Viral Load.

**Figure A3:**
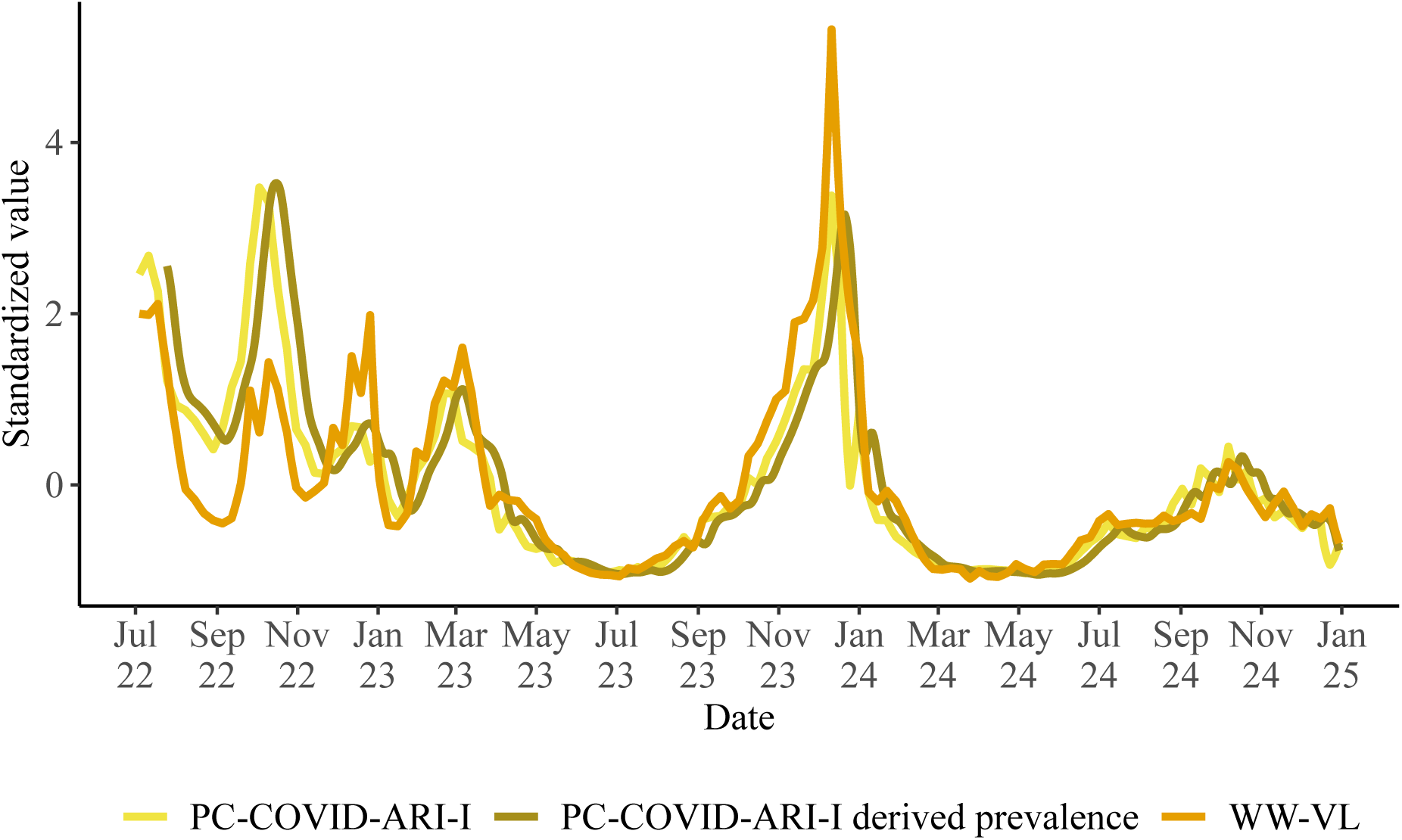
Hypothetical prevalence for PC-COVID-ARI-I. PC-COVID-ARI-I incidences were multiplied with the fecal shedding distribution shown in Figure 3 to derive a hypothetical prevalence curve. Shown are standardized values for the resulting hypothetical prevalence, the original incidence for PC-COVID-ARI and the viral load in wastewater. PC-COVID-ARI-I = Sentinel for Electronic Recording of Diagnoses of Acute Respiratory Infections, combined with Participatory System, WW-VL = Wastewater Viral Load.

**Figure A4:**
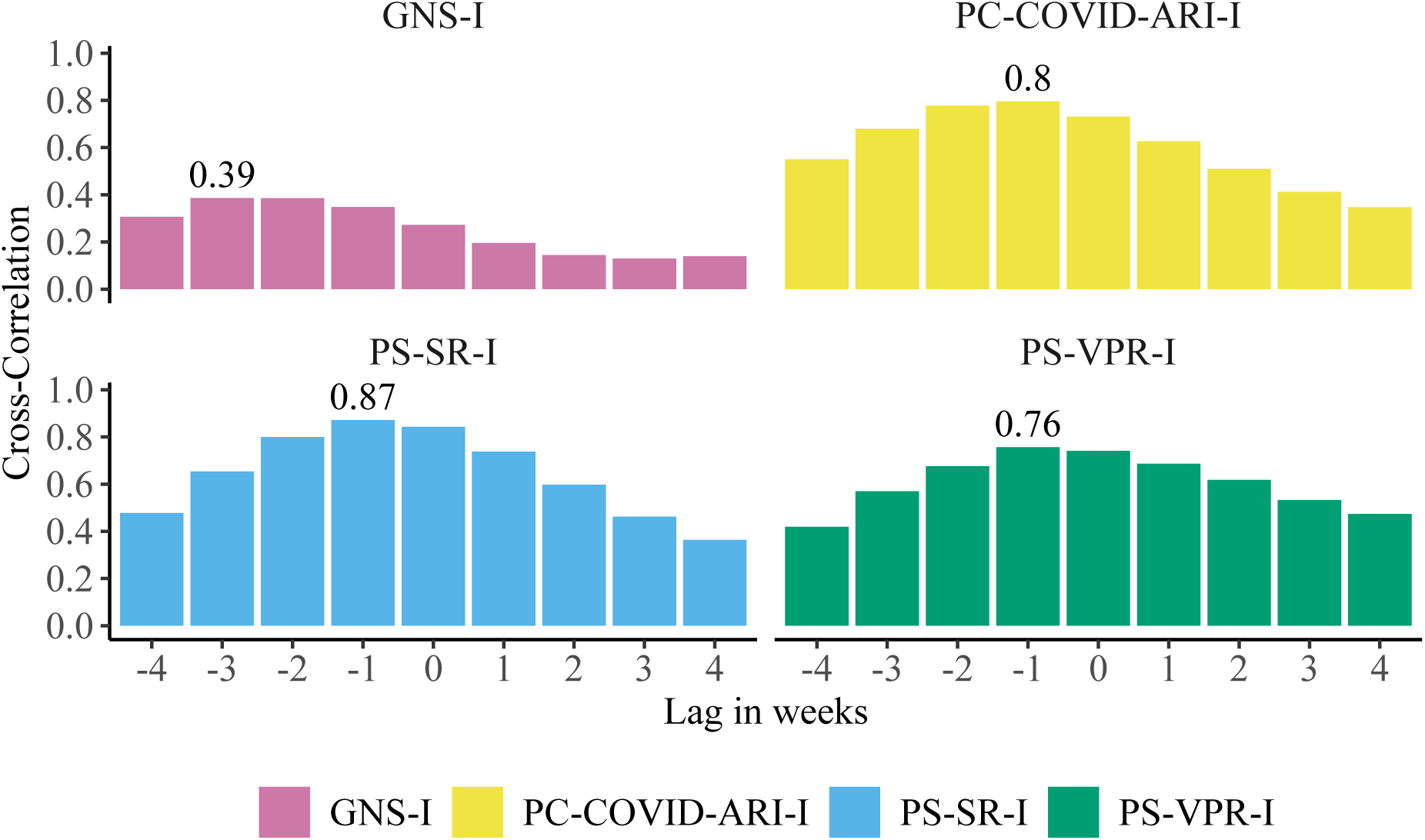
Cross-correlation between wastewater viral load and hypothetical prevalence derived from four COVID-19 incidence indicators at varying lags in weeks. Incidences were multiplied with the fecal shedding distribution shown in Figure 3 to derive a hypothetical prevalence curve for the respective incidence indicator. A correlation for a lag of +2 weeks, e.g., indicates the correlation between current prevalence and wastewater viral load measured two weeks back in time. GNS-I = German Notification System of COVID-19, PC-COVID-ARI-I = Sentinel for Electronic Recording of Diagnoses of Acute Respiratory Infections, combined with Participatory System, PS-SR-I = Participatory System Self-Reported, PS-VPR-I = Participatory System Virological Positivity Rate.

